# Validation of a physics-based computational model of epicardial and microvascular coronary physiology against continuous infusion thermodilution

**DOI:** 10.1101/2025.11.09.25339841

**Authors:** Daniel J Taylor, Tom Newman, Karolina Tlałka, Eron Yones, Kenneth Anigboro, Harry Saxton, Xu Xu, Alberto Biancardi, Rebecca Gosling, Krzysztof Czechowicz, Andrew Narracott, Rod Hose, Dominika Ciupek, Alessandro Maino, Federico Marin, Samer Fawaz, Ian Halliday, OxAMI Investigators, Julian P Gunn, Giovanni Luigi De Maria, Thomas R Keeble, Paul D Morris

## Abstract

**Introduction:** Coronary microvascular dysfunction is associated with myocardial ischaemia and an adverse prognosis, but accurate invasive assessment is technically challenging, requires dedicated hardware and increases procedure time and cost. Assessment can be performed with three-dimensional (3D) computational fluid dynamics (CFD) modelling, but this technique lacks robust in-vivo validation. In this study, we compared the accuracy of a 3D CFD method against continuous thermodilution.

**Methods:** Patients with acute and chronic coronary syndromes, undergoing continuous thermodilution and angiography-derived assessment, were recruited from two tertiary cardiac centres. Microvascular resistance reserve (MRR) and absolute flow were computed using 3D CFD in reconstructed coronary arteries. Invasive pressure measurements informed the CFD boundary conditions.

**Results:** Paired flow results were available for 131 arteries from 89 patients. Median computed MRR was 2.31 [1.83 – 3.00], which was significantly lower than continuous thermodilution assessed MRR (2.79 [2.21 – 3.52], z = 3.57, p = 0.0004). There was evidence of a moderate relationship between computed and measured MRR (ρ = 0.58, p<0.0001) and area under the receiver operator characteristic curve was 0.77 (95% CI 0.68–0.86), indicating fair classification. There was a modest relationship between computed and measured hyperaemic vessel inlet flow (ρ=0.44, p<0.0001). For both MRR and flow, agreement improved at lower, more clinically relevant, values.

**Discussion:** In this clinical validation study, when compared with continuous thermodilution, the novel CFD method demonstrated a moderate correlation, and fair diagnostic accuracy. This CFD method may be a useful, low-cost and less-invasive tool in the assessment of microvascular physiology that could complement current invasive techniques.

**Graphical Abstract:** 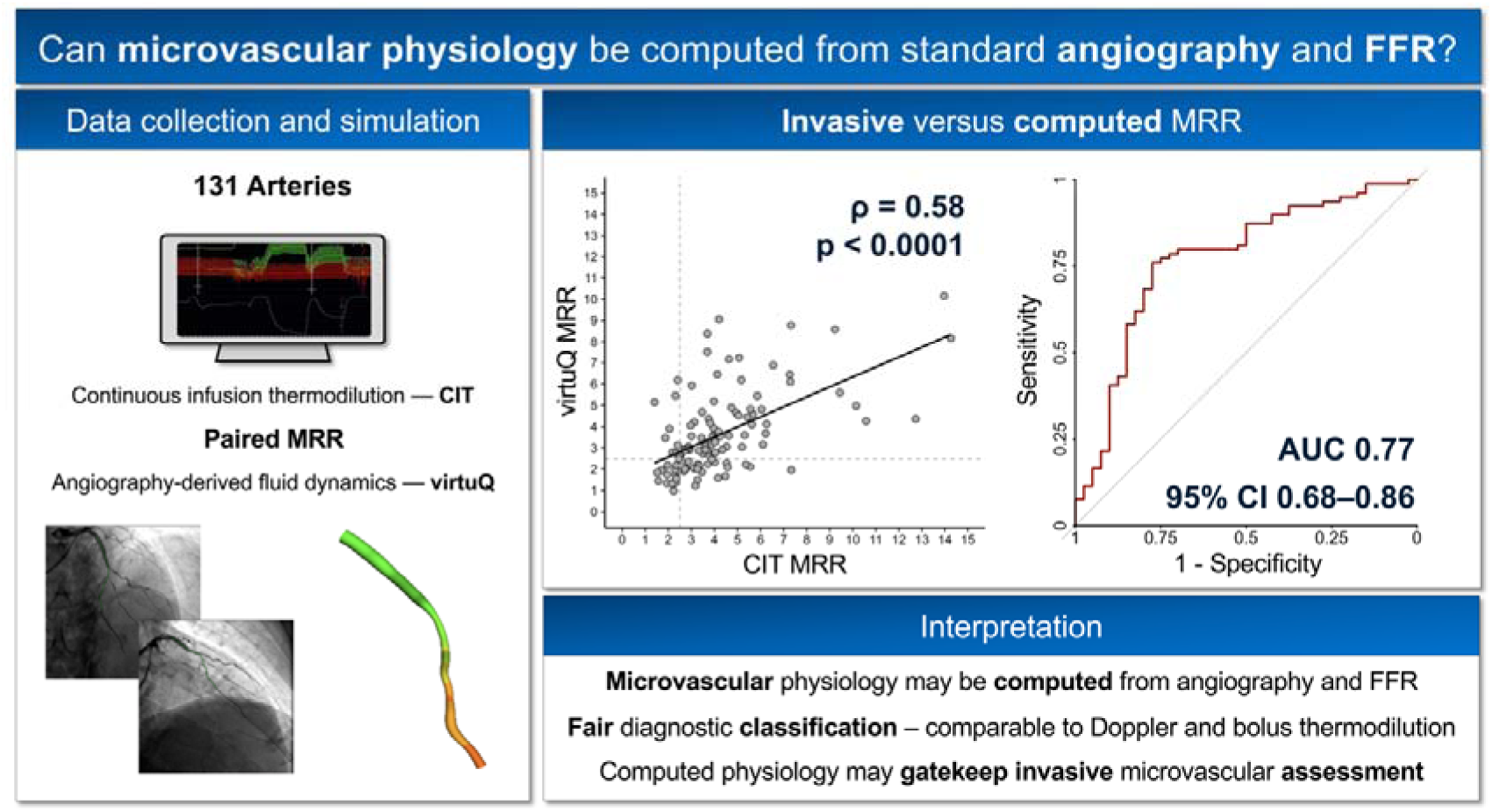

## Introduction

In the invasive management of coronary artery disease (CAD), the use of intra-coronary physiological assessment is associated with improved clinical outcomes, reduced costs and is recommended in international guideline documents (1, 2). Pressure-based assessment of fractional flow reserve (FFR) is now widely adopted, to identify ischaemia causing lesions in the epicardial coronary arteries and to guide percutaneous coronary intervention (PCI) (3). FFR however, does not yield any information about the coronary microcirculation which is increasingly implicated as a prognostically important cause of ischaemia and angina, especially in women (4). To assess the coronary microcirculation, an estimation of coronary flow must be made. This adds technical complexity, time and cost to the procedure. Common techniques for assessing microvascular physiology are bolus thermodilution and Doppler. These estimate surrogates of coronary blood flow (mean transit time and flow velocity) to derive functional indices like coronary flow reserve (CFR) and microvascular resistance reserve (MRR). Abnormalities of these surrogates are associated with prognostically significant endpoints (5) and functional outcomes (6). Yet, correlation and agreement between these techniques is often poor, even in expert hands (7). Continuous infusion thermodilution provides an alternative with more robust repeatability (8) and superior accuracy (9, 10), but requires a dedicated catheter (Rayflow, Hexacath, France), and software module (Coroventis, Uppsala, Sweden) which quantifies flow at the location of an infusion microcatheter which also adds to procedure complexity, time and cost.

Computational fluid dynamics (CFD) modelling has been used to assess coronary physiology during routine angiography and FFR assessment using a system called virtuQ (University of Sheffield, UK) (11, 12). Thus far, however, clinical validation is limited to *in-vitro* studies (11) and a small cohort of healthy vessels (13). Given that the CFD model was developed for use in the context of CAD, the *in-vivo* accuracy for this method remains unknown. The primary aim of this clinical validation study was to quantify the agreement of MRR between virtuQ and continuous thermodilution. Secondary aims included quantification of agreement for absolute flow at the vessel inlet, total vessel resistance and CFR along with identification of primary sources of error in the CFD workflow.

## Methods

### Patient recruitment and clinical data collection

Clinical data for this multi-centre cohort study were sourced from the Oxford Heart Centre and the Essex Cardiothoracic Centre. This included adult patients undergoing clinically indicated invasive coronary angiography and coronary physiological assessment, who were prospectively enrolled into the OxAMI (Oxford Acute Myocardial Infarction) and Essex-SAAMI (Essex Stable Angina and Acute Myocardial Infarction) studies respectively. Data collection for research purposes was approved by Regional Ethics Committees (OxAMI REC number 10/H0408/24, Essex-SAAMI REC reference 22/EE/0016), compliant with the Declaration of Helsinki and all patients gave written informed consent. Patients presenting with either acute (ST-elevation myocardial infarction (STEMI), non-ST elevation myocardial infarction (NSTEMI)) or chronic coronary syndromes (CCS) were considered suitable for inclusion. Exclusion criteria included severe renal impairment (estimated glomerular filtration rate <30 ml/min), contraindication to the induction of hyperaemia, hemodynamic or electrical instability, pregnancy or inability to consent.

Coronary angiograms were acquired following standard clinical protocols, using a 6 French guide catheter, with images optimised for computational reconstruction. A thermo-pressure sensitive guidewire (PressureWire X, Abbott Vascular, Santa Clara, California) was placed in the guide catheter tip, the pressure equalised, and subsequently advanced into the distal coronary artery. The dedicated, continuous thermodilution Rayflow microcatheter (Hexacath, Paris, France) was advanced over the guidewire and absolute coronary flow (mL/min), under both baseline and saline-induced hyperaemic conditions, assessed using the Coroventis (Coroflow, Uppsala, Sweden) system (14). The 0.84mm diameter continuous thermodilution microcatheter, measures absolute coronary flow by continuously infusing room temperature saline into the proximal coronary artery and computes flow at the location of infusion from downstream temperature dynamics. The continuous thermodilution method therefore dependent upon complete mixing of blood and saline infusate upstream to distal temperature measurement, which has been investigated in previous computational modelling studies (15). In addition to absolute flow, MRR, vessel resistance distal to the continuous thermodilution microcatheter outlet and CFR were calculated using:

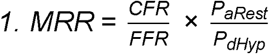

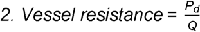

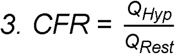

where Q represents flow, while P_a_ and P_d_ denote aortic and distal coronary pressure respectively. The subscripts *_Hyp_* and *_Rest_* correspond to measurements taken under saline-induced hyperaemia and resting conditions respectively. Pseudonymised angiography (DICOM) and physiological data were exported to the University of Sheffield for offline computational processing and statistical analysis.

### Coronary reconstruction and flow simulation

The CFD workflow (virtuQ), including vessel reconstruction protocol, has been described previously (11, 16). In brief, two angiographic projections, acquired ≥ 30° apart, displaying the vessel of interest were selected. Cases without two projections clearly displaying the vessel and lesion(s) of interest or in surgically grafted vessels were excluded. End-diastole was selected manually, guided by the ECG, and table movement artefact was corrected for. The inlet and outlet of the arterial reconstruction were selected by the operator, corresponding to the location of the continuous thermodilution infusion head (i.e., location of invasive flow measurement) and distal pressure measurement respectively. The vessel centreline and borders were traced semi-automatically from the imaged contrast gradient, with manual correction if required. Finally, a rigid, locally axisymmetric 3D reconstruction was generated automatically using an epipolar line method (16). Anatomical reconstruction accuracy was verified by an interventional cardiologist against the original angiogram. All reconstructions and physiological simulations were performed blinded to invasive physiology.

Absolute coronary flow, during hyperaemia, was then computed from a CFD simulation using P_a_ and P_d_ values as static boundary conditions at the vessel inlet and outlet respectively. To account for side branch losses, taper of the reconstructed main vessel was used to infer branch magnitude and flow loss using the Huo-Kassab law (17):

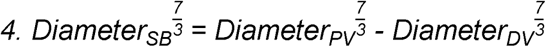

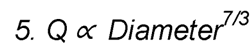

where PV and DV denote diameter of the reconstructed (main) vessel proximal and distal to the side branch (SB). The appropriateness of this scaling law for human coronary arteries has been previously demonstrated (18). In simulations, side branch flow losses were distributed continuously along the vessel length and were removed by ‘leaking’ flow through a porous arterial wall (12). All simulations used standard parameters for blood viscosity μ = 0.0035 Pa.s and density ρ = 1050 kg/m^3^.

### Statistical analysis

Categorical variables are presented as frequency (percentage). Continuous variables are presented as mean ±standard deviation or median [interquartile range] for normally distributed and skewed data respectively. Normality of data distribution was assessed using histograms and, where inconclusive, the Shapiro–Wilk test. Mean values were compared using t-tests and the Wilcoxon signed-rank test as appropriate. The relationship between haemodynamic variables was assessed with Spearman’s ρ and linear regression. Agreement was assessed with Bland Altman plots. Where data did not meet the parametric assumptions for the original Bland Altman method (19), we derived median bias and limits of agreement using quantile regression at the 50^th^, 2.5^th^ and 97.5^th^ centiles (20, 21). The diagnostic performance of CFR (prespecified threshold ≤2.5 (22)) and MRR (prespecified threshold ≤2.5 (23)) were quantified with receiver operator characteristic (ROC) curves and associated area under the curve (AUC) values computed using DeLong’s method. We calculated diagnostic accuracy, sensitivity, specificity, positive predictive value (PPV) and negative predictive value (NPV) with associated 95% CI computed using the Clopper-Pearson exact method and Locally Estimated Scatterplot Smoothing (LOESS) plots to visually display classification concordance. All statistical tests were two-tailed unless otherwise specified. Analyses were performed using RStudio version 2024.04.2+764, using the pROC, binom.test and ggplot2 packages.

We also performed an error propagation analysis to assess predictors of agreement between continuous thermodilution-and CFD-derived flow and MRR. This hypothesis-driven secondary analysis considered 13 variables likely to influence the physics underpinning CFD simulations and not demographic/comorbid data likely to be associated with coronary microvascular disease (CMD). Details of included variables, along with rationale, are given in the supplementary material. As agreement was unlikely to be linearly related with multiple variables (e.g., at the extremes of FFR, we expected agreement to be worse), we used regression and SHapley Additive exPlanations (SHAP) analysis to determine the variables most influential for agreement for the corresponding machine learning model explaining large portion of a variance in presented dataset. SHAP results are presented as a dimensionless value, with larger numbers denoting a greater overall contribution to model agreement. See supplementary material for further methodological details.

## Results

### Study population

One hundred and two patients were included, in whom invasive physiology was assessed in 178 arteries. Thirteen patients and 43 vessels were excluded, primarily due to insufficient angiographic views for arterial reconstruction, leaving 89 patients and 135 vessels in the final analysis (see supplementary material for exclusion flowchart and characteristics of excluded patients and vessels). Of the included patients, 70 (79%) were male and mean age was 63 ±11 years. The left anterior descending artery (LAD) was the most frequently included target vessel (n = 83 (62%)) and the median FFR was 0.81 [0.70 – 0.89] (see supplementary for full invasive FFR data). Of the reconstructed vessels, 108 (81%) contained an epicardial lesion with >10% 3D quantitative coronary angiography (QCA) diameter stenosis. Most lesions (n = 87 (81%)) were in the proximal – mid vessel and median diameter stenosis across all cases was 31% [19 – 47] (see table 1 for demographics and vessel characteristics and supplementary material for lesion data including screenshots of every reconstructed vessel).

**Table 1:** Patient demographics and lesion characteristics of included cases. BMI, body mass index; CCS, chronic coronary syndrome; NSTEMI, non-ST elevation myocardial infarction; PCI, percutaneous coronary intervention; QCA, quantitative coronary angiography; STEMI, ST-elevation myocardial infarction.

| Demographics (n = 89) |  |
| --- | --- |
| Age, y | 63.5 ±10.6 |
| Male | 70 (79%) |
| BMI | 30.3 ±8.8 |
| Current or ex-smoker | 47 (53%) |
| Enrolment pathology |  |
| CCS | 32 (36%) |
| STEMI | 19 (21%) |
| NSTEMI | 38 (43%) |
| Comorbidities |  |
| Left ventricular ejection fraction | 52.1 ±7.2 |
| Hypertension | 38 (43%) |
| Diabetes mellitus | 18 (20%) |
| Hypercholesterolaemia | 36 (40%) |
| Previous myocardial infarction | 24 (27%) |
| Previous stroke | 47 (53%) |
| Vessel characteristics (n = 135) |  |
| Left anterior descending | 82 (62%) |
| Left circumflex | 13 (10%) |
| Right coronary artery | 33 (25%) |
| Diagonal branch | 2 (1%) |
| Obtuse marginal | 2 (1%) |
| Intermediate | 1 (1%) |
| Pre-PCI physiology assessment | 64 (48%) |
| Vessel inlet diameter (mm) | 3.0 ±0.5 |
| Vessel outlet diameter (mm) | 1.9 [1.7 – 2.2] |
| Lesion characteristics |  |
| Stenosis present | 108 (81%) |
| 3D-QCA percentage stenosis | 31 [19 – 47] |
| Stenosis location |  |
| Proximal | 44 (41%) |
| Mid | 43 (40%) |
| Distal | 21 (20%) |
| Tandem lesions | 19 (18%) |
| Diffuse disease | 31 (29%) |
| FFR | 0.81 [0.70 – 0.89] |
| FFR significant disease (≤0.80) | 62 (47%) |

### MRR

MRR was computed successfully in 116 vessels. Median computed MRR was 2.31 [1.83 – 3.00], which was significantly lower than median continuous thermodilution-assessed MRR (2.79 [2.21 – 3.52], z = 3.57, p = 0.0004). There was evidence of a moderate relationship between computed and continuous thermodilution-assessed MRR (ρ = 0.58, p < 0.0001). Across all cases, there was a tendency for virtuQ to underestimate continuous thermodilution-assessed MRR, but with improved agreement at lower values, closer to the diagnostic threshold (figure 1). Overall diagnostic accuracy of computed MRR was 63.0% (95% CI 53.7 – 71.7) and AUC was 0.77 (95% CI 0.68 – 0.86), indicating fair diagnostic classification (table 2, figure 2). In keeping with the predefined diagnostic threshold of 2.5, ROC sensitivity analysis identified an optimal diagnostic threshold for computed MRR of 2.5 (supplementary).

**Figure 1.**
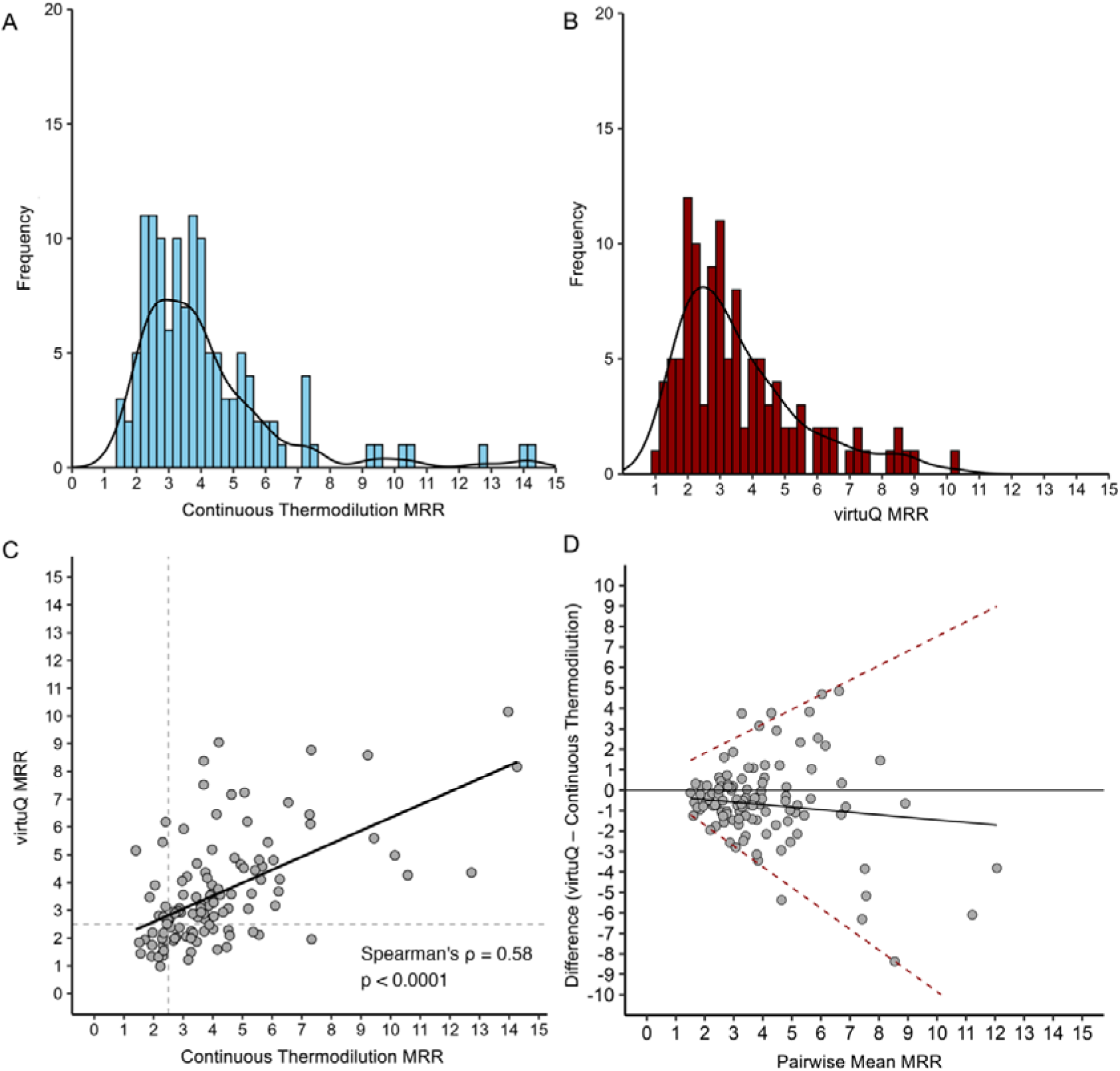
Summary and agreement results for MRR. A) Histogram of continuous thermodilution-assessed MRR. B) Histogram of computed (virtuQ) MRR. C) Correlation plot and simple least regression for continuous thermodilution versus virtuQ derived MRR. D) Bland Altman plot demonstrating agreement between continuous thermodilution and virtuQ derived MRR. Median bias (solid line) and 95% limits of agreement (dashed lines) derived from quantile regression. MRR, microvascular resistance reserve.

**Figure 2.**
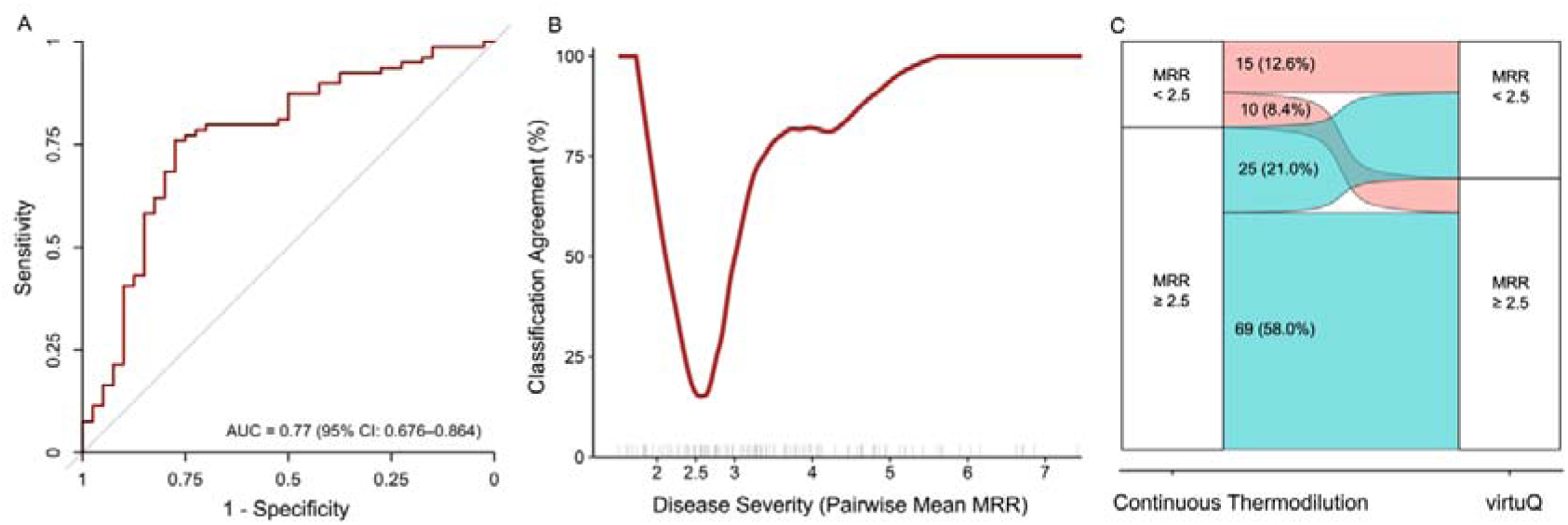
Diagnostic accuracy results for MRR. A) Receiver operator characteristic curve for MRR using a diagnostic threshold of 2.5. B) LOESS-smoothed curve showing the percentage agreement between virtuQ and Rayflow MRR classifications across the range of disease severity, with individual data points indicated by rugs along the x-axis. C) Alluvial plot at diagnostic threshold 2.5. AUC, area under the curve; MRR, microvascular resistance reserve.

**Table 2:** Summary diagnostic accuracy results for total vessel resistance, microvascular resistance reserve (MRR) and coronary flow reserve (CFR). Median bias and 95% LOA computed with Bland-Altman quantile regression plots. 95% CI for sensitivity, specificity, positive and negative predictive values calculated with the Clopper–Pearson exact confidence interval and AUC 95% CI calculated with DeLong’s method. AUC, area under the curve; LOA, limits of agreement.

|  | MRR | CFR |
| --- | --- | --- |
| Successful simulations | 116 (85.9%) | 116 (85.9%) |
| Median Bias at diagnostic threshold | -0.61 | 0.04 |
| 95% LOA at diagnostic threshold | -2.23 to 1.67 | -2.54 to 1.88 |
| Accuracy, % (95% CI) | 63% (54 – 72) | 72% (62 – 80) |
| Sensitivity, % (95% CI) | 52% (41 – 63) | 23% (10 – 41) |
| Specificity, % (95% CI) | 85% (70 – 94) | 89% (81 – 95) |
| Positive predictive value, % (95% CI) | 87% (74 – 95) | 44% (20 – 70) |
| Negative predictive value, % (95% CI) | 47% (35 – 59) | 76% (66 – 84) |
| AUC (95% CI) | 0.77 (0.68 – 0.86) | 0.69 (0.58 – 0.80) |

### Absolute flow

Hyperaemic flow was computed successfully in 131 vessels. At the vessel inlet, corresponding to the location of continuous thermodilution assessment, median computed hyperaemic flow was 194 [127 – 272] mL/min. This was significantly larger than continuous thermodilution-assessed flow (median 157 [112 – 205] mL/min, z = - 4.5915, p < 0.0001). There was evidence of a moderate relationship between computed and continuous thermodilution-assessed hyperaemic flow (ρ = 0.44, p < 0.0001). In arteries drawing lower flow (typically <200 mL/min), there was minimal overall bias between continuous thermodilution-assessed and computed flow. However, at higher flow values, computed flow significantly overestimated continuous thermodilution-assessed flow with an associated worsening of limits of agreement (figure 3).

**Figure 3.**
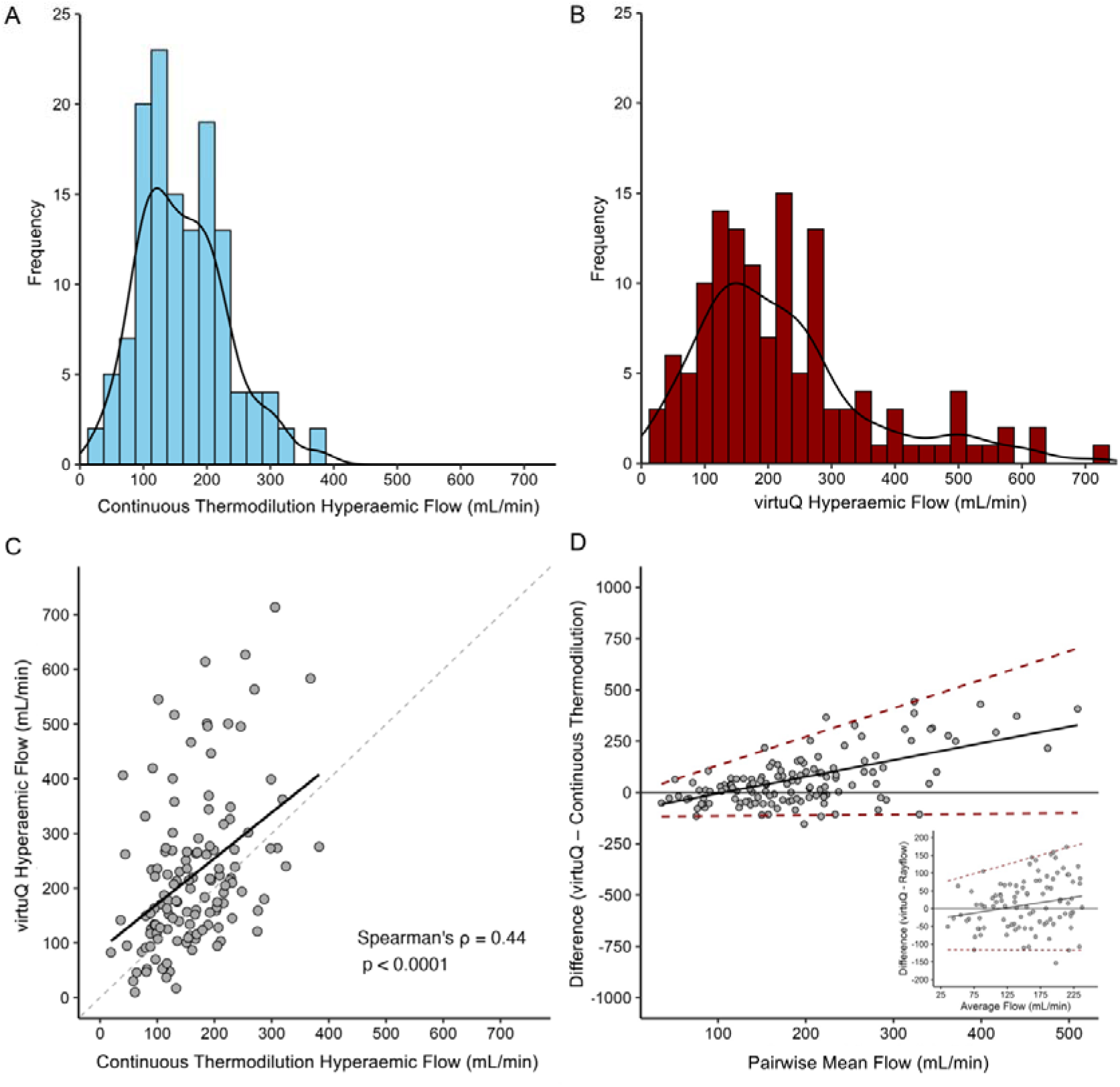
Summary results for hyperaemic flow. A) Histogram of continuous thermodilution-assessed hyperaemic flow. B) Histogram of computed (virtuQ) hyperaemic flow. C) Correlation plot and simple least regression for continuous thermodilution versus virtuQ derived hyperaemic flow. D) Bland Altman plot demonstrating agreement between continuous thermodilution and virtuQ derived hyperaemic flow. Median bias (solid line) and 95% limits of agreement (dashed lines) derived from quantile regression. The sub figure shows a magnified Bland Altman plot at the lowest flow values.

### Distal vessel resistance and CFR

Hyperaemic vessel resistance was computed successfully in 131 vessels. Median computed vessel resistance was 360 [238 – 524] WU, which was significantly lower than continuous thermodilution-assessed resistance (median 452 [350 – 586] WU, z = -2.82, p = 0.0048). There was evidence of a weak relationship between computed and continuous thermodilution-assessed vessel resistance (ρ = 0.39, p < 0.0001). For typical cases below 500 WU, there was a tendency for virtuQ to overestimate continuous thermodilution-assessed resistance and vice-versa above this value (supplementary). CFR was computed in 116 vessels, disclosing a median value of 1.57 [1.33 – 1.94]. Computed CFR was significantly lower than median continuous thermodilution-assessed CFR (1.98 [1.60 – 2.55], z = -3.43, p = 0.0006), with evidence of a weak relationship between these values (ρ = 0.36, p < 0.0001). For cases with a CFR below the diagnostic threshold, there was a tendency for virtuQ to overestimate continuous thermodilution-assessed CFR and vice-versa above the diagnostic threshold. Across all cases, agreement was improved at lower CFR values (supplementary). Overall diagnostic accuracy of computed CFR was 71.6% (95% CI 62.4 – 79.5) and AUC was 0.69 (95% CI 0.58 – 0.80)(see table 2 for full results). ROC sensitivity analysis identified an optimal diagnostic threshold for computed CFR of 2.7 (supplementary).

### Determinants of agreement

Correlation analysis identified uncertainty in the flow diameter scaling exponent as the most significant determinant of agreement for MRR (ρ = -0.36, p < 0.0001). The correlation matrix suggested no other variables, independent of side branch flow, were predictive of MRR agreement (supplementary). SHAP coefficients could not be calculated for MRR agreement, due to inability to reliably fit a predictive model (supplementary). For absolute flow, uncertainty in the flow diameter scaling (Huo-Kassab) exponent was identified as the dominant source of error (SHAP value = 72.6, ρ = -0.60, p < 0.0001), followed by FFR (SHAP value = 7.7, ρ = 0.244, p = 0.0051). A third variable, mismatch between FFR and CFD-derived hyperaemic stenosis resistance (HSR) was also associated with agreement (ρ = -0.40, p < 0.0001). Increasing HSR/FFR mismatch values are suggestive of either pressure losses proximal to the continuous thermodilution infusion head or inaccurate reconstruction anatomy, but this mismatch variable was excluded from SHAP analysis to avoid data overfitting (supplementary).

## Discussion

In this study, we evaluated the agreement between CFD-(virtuQ) and continuous thermodilution-derived coronary physiology. The main results were:

1. Computed MRR demonstrated a moderate relationship between continuous thermodilution-measured values with fair diagnostic classification and stronger agreement at lower, more clinically relevant, values.
2. Computed absolute hyperaemic flow demonstrated a moderate relationship with continuous thermodilution-measurements, agreement again improved at lower values.

### Current accuracy and future CFD developments

Previous validation studies of invasive techniques for quantifying MVD have shown mixed results (7, 22, 24–26). The largest study that compared bolus thermodilution-against Doppler-derived index of microvascular resistance, in two expert centres, revealed only a modest correlation between the two methods measuring the same index (r = 0.43; P < 0.0001)(7). More recent data from 175 patients demonstrated a lower correlation coefficient between bolus and continuous thermodilution of 0.26 (22), and further work suggesting this may be as low as 0.11 (27, 28). In the current study, CFD-derived MRR was the most directly applicable metric for quantifying microvascular-specific physiology (i.e., independent of epicardial lesion severity). Our reported correlation between CFD and continuous thermodilution-measured MRR (0.58, p < 0.0001) is therefore, relatively strong in comparison to previous work. In addition, agreement was strongest at the lower, more clinically relevant, values around the diagnostic threshold. Thus, the current study suggests that, with continuous thermodilution as the reference, the accuracy of the novel CFD method is at least comparable to routinely used techniques like Doppler and bolus thermodilution in terms of assessing microvascular physiology. The technique may be implemented in a standard catheterisation laboratory, with any pressure-sensitive guidewire, to produce results within five minutes. Furthermore, in the current study, the relationship between continuous thermodilution-assessed and CFD-derived MRR was either superior (29) or comparable (30–32) with that reported for other angiography-derived (computed) techniques for microvascular assessment.

The relationship between computed and continuous thermodilution-assessed absolute flow at the vessel inlet was moderate. Error propagation analysis suggested that error in the CFD method was attributable primarily to the method of side branch flow incorporated into the CFD simulation. This is consistent with similar previous analyses (33). For the current study, the side branch method used a population-averaged scaling exponent of 7/3, supported by theoretical (17) and meta-analysis studies (18), but further work may wish to investigate novel models of side branch flow incorporation to reduce error (34). Additional sources of error included mismatch between FFR and CFD-derived HSR. This may be attributable to either the artificial stenosis created by the continuous thermodilution microcatheter in the left main stem, organic causes of pressure loss proximal to the microcatheter infusion head (i.e., left main stem disease) or inaccuracies in 3D reconstructed anatomy. Agreement was also modestly dependent on FFR, which may be multifactorial in aetiology; a low pressure gradient principally sensitises the simulation to errors in reconstruction diameter. Additionally, with minimal epicardial stenoses, dynamic pressure losses within the vessel become non-negligible and these are not currently accounted for in CFD simulations. These factors may therefore provide targets to improve the CFD workflow and adjust clinical data collection protocol for further prospective validation against continuous thermodilution.

### Clinical implications

The importance of MVD is increasingly recognised (1, 2) and the subject of ongoing research (35, 36). In the context of CCS, current guidelines advocate for assessment of MVD in the absence of significant (obstructive) epicardial disease (1). Indeed, a seminal controlled trial of MVD treatment (CorMicA) randomised only patients without obstructed coronary arteries (37). Given the increased cost and time associated with invasive microvascular assessment, this appears a sensible approach, because it strikes a balance between test sensitivity and the pressures of real-world clinical practice. However, the assumption of a predominant phenotype where pathology lies exclusively in the microvascular compartment appears biologically unlikely; large registry data has shown MRR to be an independent predictor of major adverse cardiac events (MACE) in patients with obstructive epicardial stenoses (5). Further, recent re-analysis of the ORBITA trial (38), using virtuQ, suggested MVD may mediate the symptomatic response to PCI in the context of flow-limiting epicardial stenoses despite optimal medical therapy (39). In the context of acute coronary syndromes, the indication for invasive physiological assessment is less clear (IIb, B recommendation)(40). However, as MVD is independently predictive of MACE (41), its assessment may help stratify those in need of aggressive pharmacotherapy. Our CFD method may therefore address this unmet need, allowing quantification of MVD in a wide cohort of patients using routinely available clinical data.

Should focused microvascular assessment be considered for a wider patient cohort, our results suggest virtuQ may effectively rule MVD in or out in a majority of patients and identify those most likely to benefit from full invasive assessment. Taking the results of this study, using MRR thresholds of 1.8 and 2.5 for ruling microvascular resistance in or out respectively would include 89 (76.7%) vessels and yield sensitivity and specificity of 87.3% and 85.0% respectively. As total vessel resistance and CFR are influenced by disease in both the microvascular and epicardial compartments, their specificity for MVD detection in stenotic vessels is limited. MRR may therefore provide the most universally applicable microvascular assessment. It is also reassuring that the sensitivity analyses identified a diagnostic threshold for MRR (2.5) which is consistent with the results of other invasive studies and the guidelines (23, 26).

### Limitations

None of the current invasive techniques for measuring flow or microvascular resistance represent a ‘ground-truth’, and this should be borne in mind when interpreting the results. Absolute resistance measurements in the distal third of the vessel are unreliable with continuous infusion, due to the artificial stenosis created by the microcatheter, meaning direct validation of virtuQ absolute resistance at the vessel outlet is not possible. The virtuQ technique requires a distal pressure measurement, necessitating coronary instrumentation. Case selection is known to influence accuracy in validation studies (42), and while we have endeavoured to fully describe our current dataset, ability to draw conclusions about comparability with other techniques is limited. As pilot validation data suggested agreement between virtuQ and continuous thermodilution did not meet the assumptions required for simple Bland-Altman analysis (13), a power calculation for sample size could not be calculated (43). Agreement data for numerically higher flow and resistance values are derived from a relatively small number of data points and lack statistical power.

Furthermore, accuracy may be poorer in patients undergoing assessment for angina with non-obstructive coronary arteries (ANOCA). The current study evaluated agreement only in the context of structural microvascular disease, although the same principles may be applied with pressures acquired during acute vasoreactivity testing. There is no diagnostic threshold for absolute flow (mL/min) reduction, which is the subject of ongoing research (44). As an exploratory secondary analysis, results from the SHAP analysis are hypothesis generating and should be interpreted with caution.

## Conclusion

Our CFD workflow allows simultaneous assessment of absolute coronary blood flow and indices of coronary physiology, from standard pressure wire data, with a modest agreement against continuous thermodilution measurements. MRR demonstrated the strongest association with invasive measurements, giving fair diagnostic classification and, with future work, may represent an effective strategy for gatekeeping invasive microvascular assessment.

## Data Availability

Available on reasonable request

