## Supplementary for "Validation of a physics-based computational model of epicardial and microvascular coronary physiology against continuous infusion thermodilution"

**Supplementary Material for:**

**Comparison of continuous infusion thermodilution and physics-based computational modelling for assessment of coronary epicardial and microvascular physiology**

**Contents**

Page 2 – 3:	SHAP analysis included variables

Page 4:	SHAP analysis technical specifications

Page 5:	Patient exclusion flowchart

Page 6:	Distribution of invasive FFR values in included cases

Page 7:	Distribution of epicardial lesion severity in included cases

Page 8:	Distribution of computed physiology values

Page 9 – 24:	Details of every included case

Page 25:	Demographics and lesion characteristics of excluded patients

Page 26:	Split violin plot for Rayflow and virtuQ assessed MRR

Page 27:	ROC sensitivity analysis for MRR

Page 28:	Alluvial plot for MRR

Page 29:	MRR Bland-Altman quantile regression line equations

Page 30:	MRR SHAP analysis results

Page 31:	Before and after dot plot for MRR

Page 32:	Split violin plot for Rayflow and virtuQ assessed inlet flow

Page 33:	Flow Bland-Altman quantile regression line equations

Page 34 – 36:	Absolute inlet flow SHAP analysis results

Page 37:	Before and after dot plot for hyperaemic inlet flow

Page 38:	Summary agreement results for total vessel resistance

Page 39:	Summary agreement results for CFR

Page 40:	ROC sensitivity analysis for CFR

**SHAP analysis included variables**

For SHAP analysis, we considered 13 variables. Given a total of 131 vessels with hyperaemic flow results, this preserved a ratio of variables to data points of 1:10 as to try and avoid overfitting data. The variables considered are listed below along with a brief justification:

1. Fractional flow reserve (FFR): For high FFR cases, the total pressure delta driving flow from the vessel inlet to outlet will be low. Given FFR is frequently expressed as the ratio of integer values, the precision of the pressure wire will contribute significantly greater uncertainty for health cases. Additionally, the CFD methodology does not consider dynamic pressure changes which are likely to exert a greater influence at lower pressure deltas. Finally, at very low FFR values, there is likely to be a considerable stenosis restricting adequate opacification of the vessel lumen on angiography.
2. Flow diameter scaling (Huo-Kassab) exponent: This is likely the primary determining factor when computing side branch flow (opposed to using an alternative model of leak). Given significant intra-population uncertainty has been described, we compared the differences in flow when using the Huo-Kassab exponent of 2.33 and the Murray exponent of 3.0.
3. Proximal taper: Defined as the ratio between the smallest diameter in the proximal 10% of the reconstructed vessel against the inlet diameter. A significant degree of taper in the proximal vessel may lead to unstable simulation results.
4. Proximal estasia (i.e., pathological downstream vessel widening): Defined as the ratio between the largest diameter in the proximal 10% of the reconstructed vessel against the inlet diameter. A significant degree of ectasia in the proximal vessel may lead to unstable simulation results.
5. Distal taper: Defined as the ratio between the smallest diameter in the distal 10% of the reconstructed vessel against the inlet diameter. A significant degree of taper in the distal vessel may lead to unstable simulation results.
6. Distal vessel ectasia: Defined as the ratio between the largest diameter in the distal 10% of the reconstructed vessel against the inlet diameter. A significant degree of ectasia in the distal vessel may lead to unstable simulation results.
7. Vessel length: The Rayflow technique recommends 70mm distance between the shower head and location of distal pressure measurement. A significant difference in this result, particularly lower values, are likely to influence factors such as complete mixing of blood with saline infusate and heat transfer to the vessel wall which may worsen accuracy.

8. Inlet vessel diameter: A particularly large or small inlet may be indicative of error in the vessel reconstruction. We have previously used OCT data to show the virtuQ reconstruction protocol underestimates true lumen radius in the proximal vessel.
9. Total vessel taper (inlet – outlet diameter): A high taper not only influences side branch flow but also increases pressure losses within the vessel. The balance between increasing uncertainty from side branch losses versus improved precision and simulation fidelity from pressure delta is unknown.
10. 3D quantitative coronary angiography diameter stenosis: The virtuQ technique is designed primarily for use in cases with an intermediate stenosis. In cases with a low % stenosis, there the simulation becomes increasingly sensitive to anatomical accuracy in the reconstruction inlet and outlet. Further, for very stenosed vessels, opacification of the vessel lumen on angiography may be compromised, thus limiting reconstruction accuracy.
11. Focal versus diffuse disease: The virtuQ reconstruction protocol was designed specifically for focal lesions. In cases of diffuse disease, anatomical accuracy may be worse.
12. Lesion location: Lesions in the proximal and distal vessel may not allow for sufficient development of flow within the simulation.
13. Hyperaemic stenosis (HSR)/ FFR mismatch: Computed using LOG(1 + HSR)/(1 - FFR). This provides a metric from 0 to infinity which gives an estimate of the concordance between these two measures of epicardial lesion physiology. FFR is invasively measured which HSR is computed from the reconstructed artery in virtuQ. Consequently, should a significant source of pressure loss within the vessel not be captured within the CFD simulation, mismatch will increase. Possible sources of error for a higher mismatch include significant pressure wire drift, the effect of the artificial stenosis induced by the Rayflow catheter, significant left main stem disease, inaccurate anatomical reconstruction.

**SHAP analysis technical specifications**

Before deriving SHAP coefficients, we examined for possible correlations between included variables. For absolute flow, we identified strong associations between

- FFR with 3D QCA
- Proximal taper with proximal ectasia
- Distal taper with distal ectasia
- Reconstruction inlet diameter with total vessel taper
- Flow-diameter scaling exponent with HSR/FFR mismatch

The correlation matrix is shown below

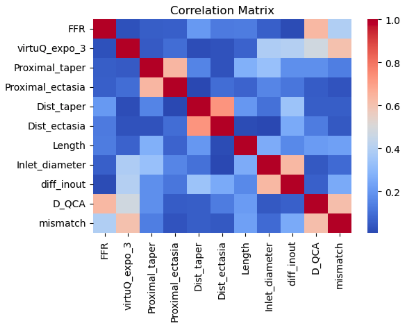

Accordingly, to reduce the likelihood of data overfitting and improve interpretability of SHAP coefficients, we removed the following variables from the analysis: 3D QCA, proximal ectasia, distal ectasia, total vessel taper and HSR/FFR mismatch.

We then split the data into training (80% of data) and test sets (20%). Random Forest regression and Ridge Linear Regression models were then trained, with the difference between computed and measured inlet flow as a target. We performed 5-fold cross validation on the training set and used the optimum model for evaluation on the test set. Because of the modest sample size and high result variability, we repeated the analysis 60 times with different train-test splits and averaged the cross-validation train R^2^ scores, the cross-validation test R^2^ scores, test R^2^ scores and SHAP values for cross-validation test folds and test set.

**Patient exclusion flowchart**

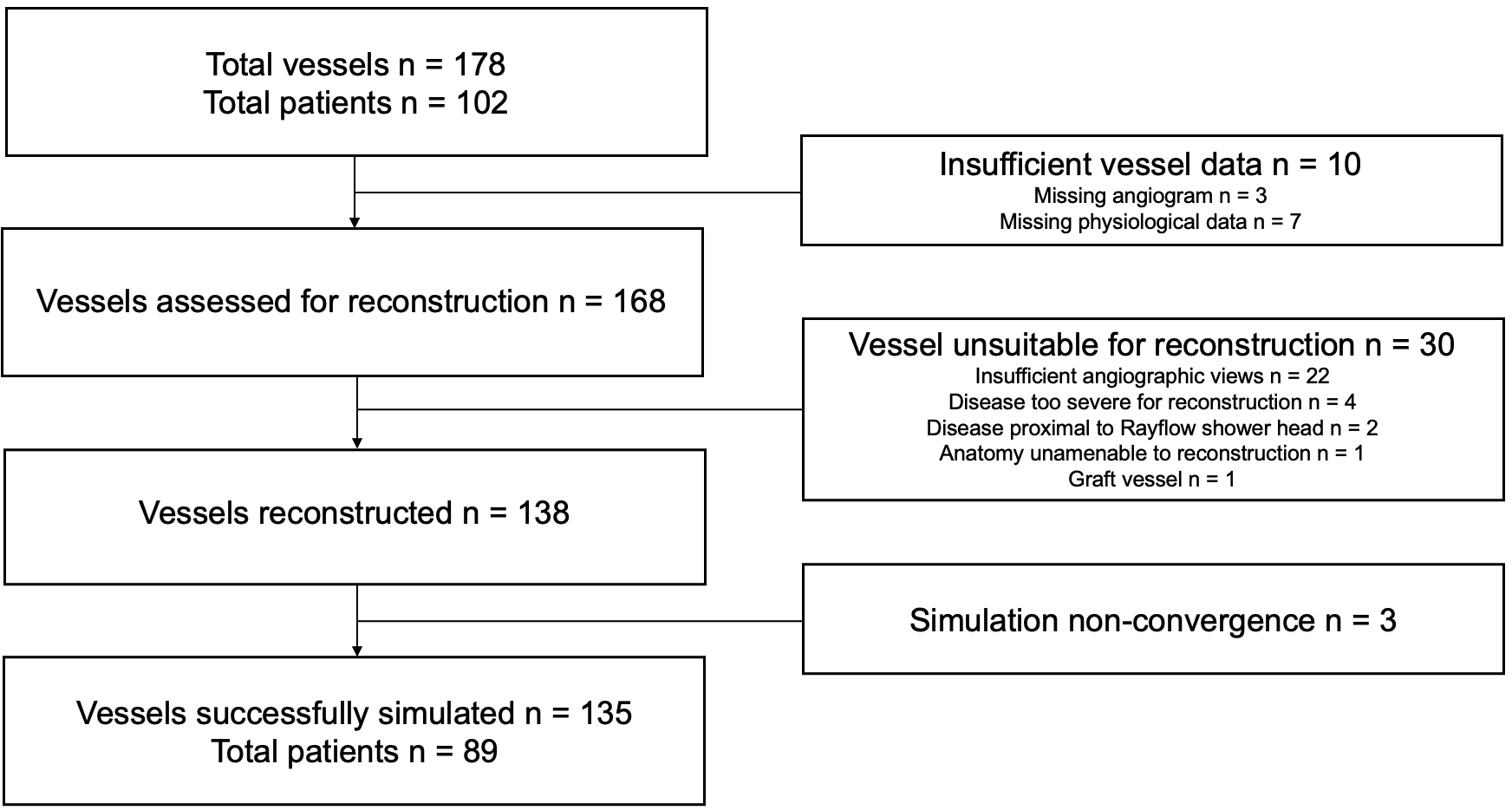

**Distribution of invasive FFR values in included cases**

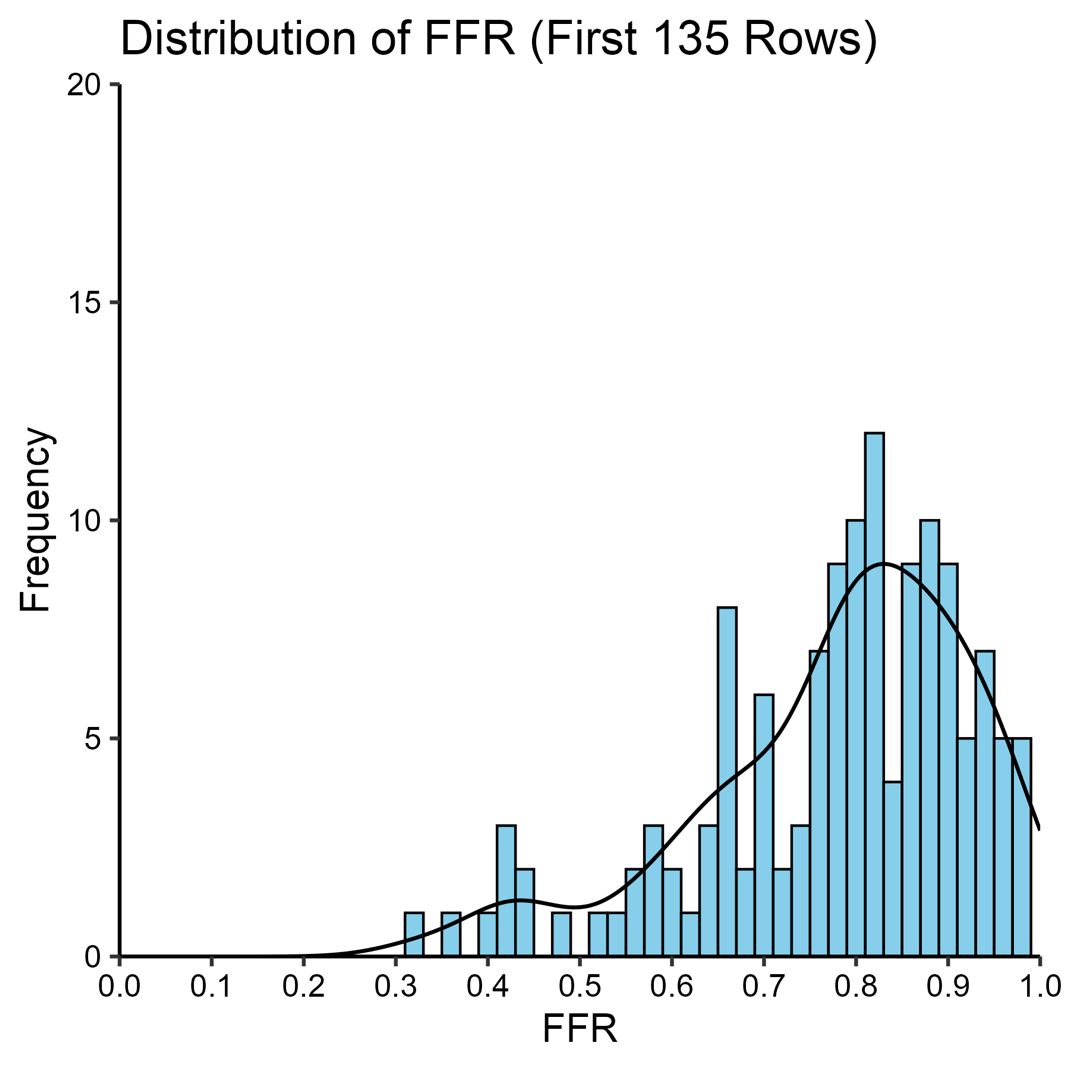

**Distribution of epicardial lesion severity in included cases**

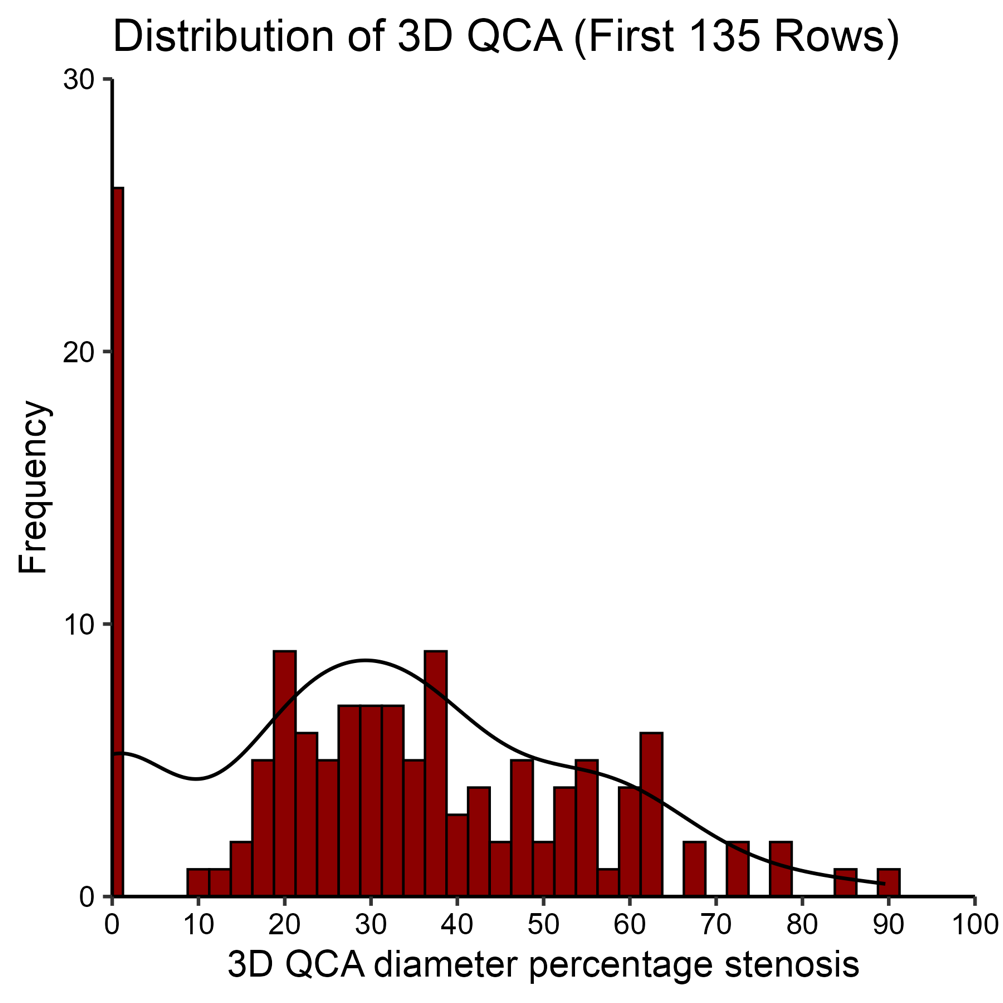

**Distribution of hyperaemic outlet flow and associated distal microvascular resistance** (MVR)(calculated from distal pressure/outlet flow) values of included cases

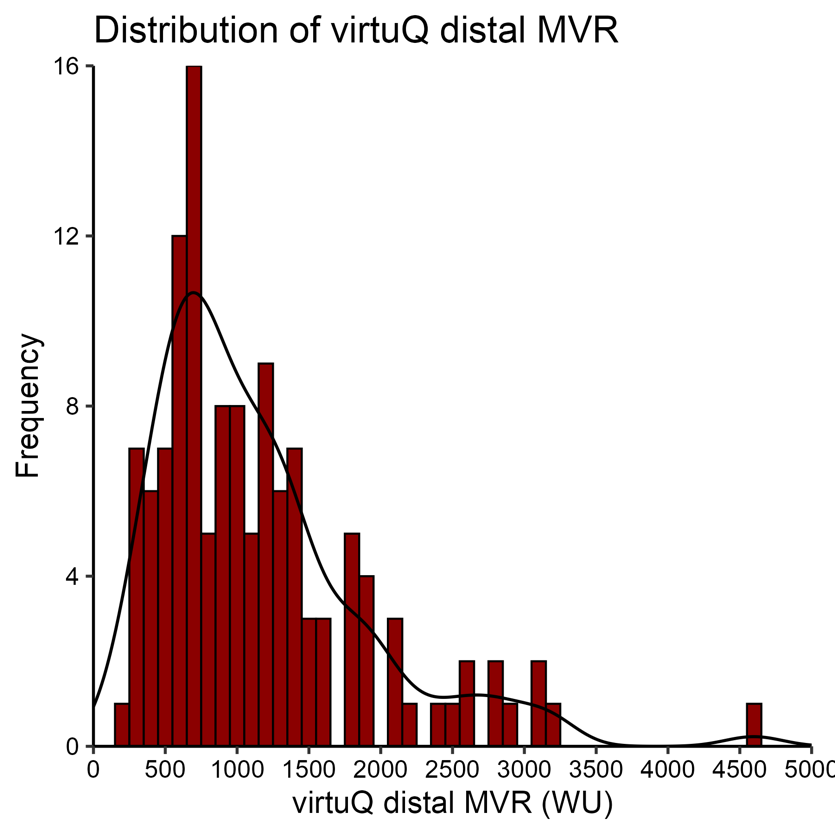

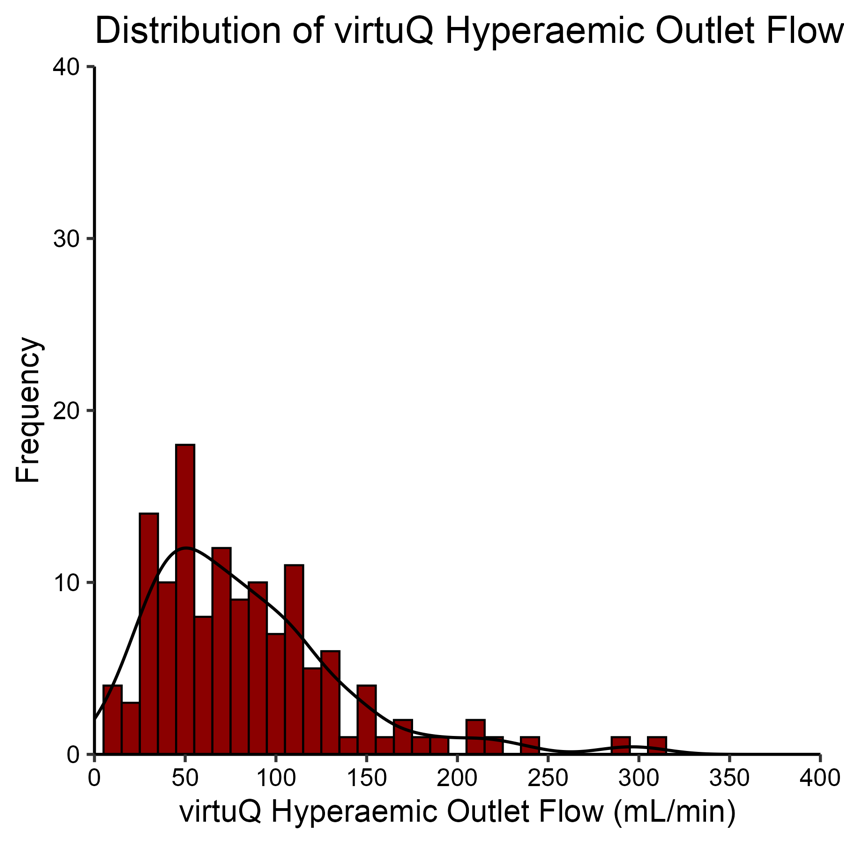

**All 131 included cases with computed hyperaemic flow.**

For every case we include a screenshot of the 3D reconstruction with pressure distribution mapped along the vessel surface, a scale for colours and corresponding pressure drop is provided below the final case. We also include angiogram screenshots with segmented vessel centreline backprojected onto the angiogram and key invasive and simulated physiology (inlet diameter, outlet diameter, 3D quantitative coronary angiography (QCA) percentage diameter stenosis, invasive FFR and simulated flow at the vessel inlet and outlet under hyperaemia).

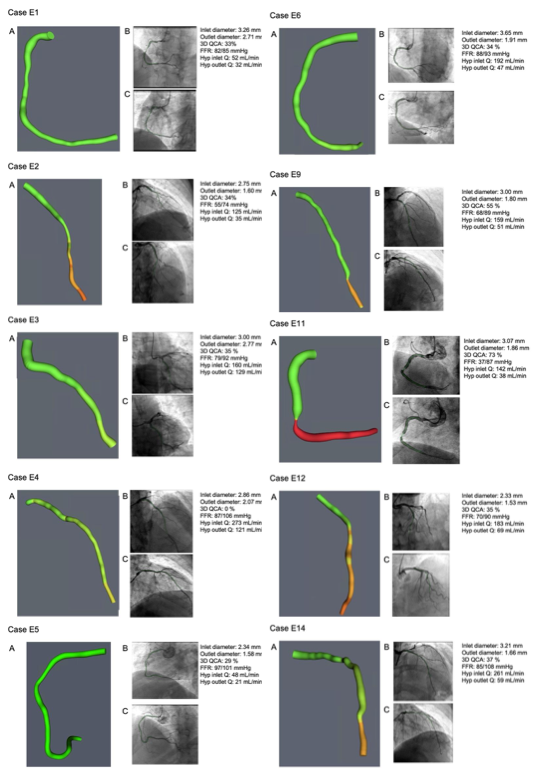

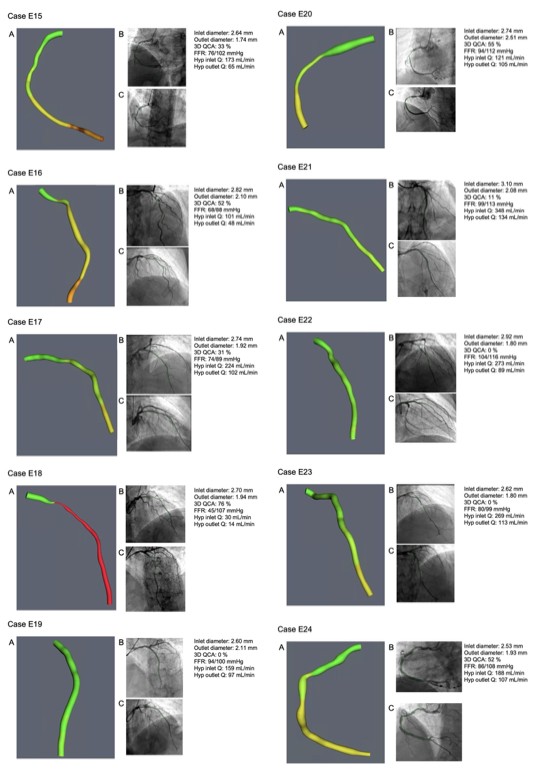

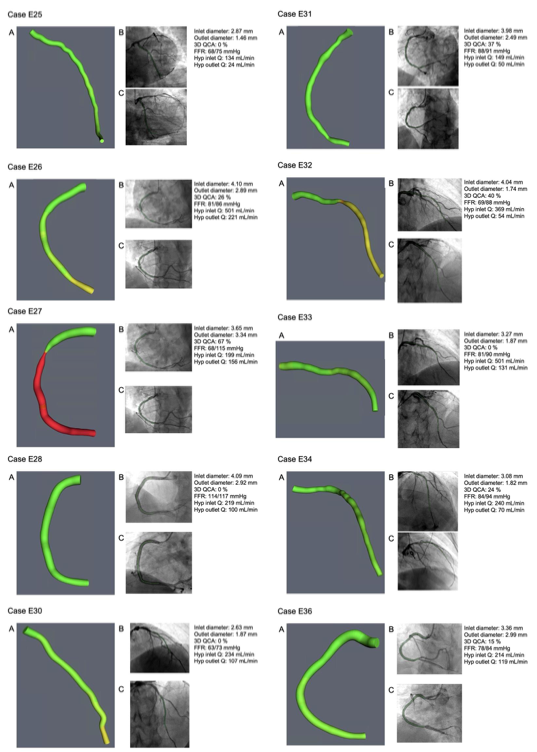

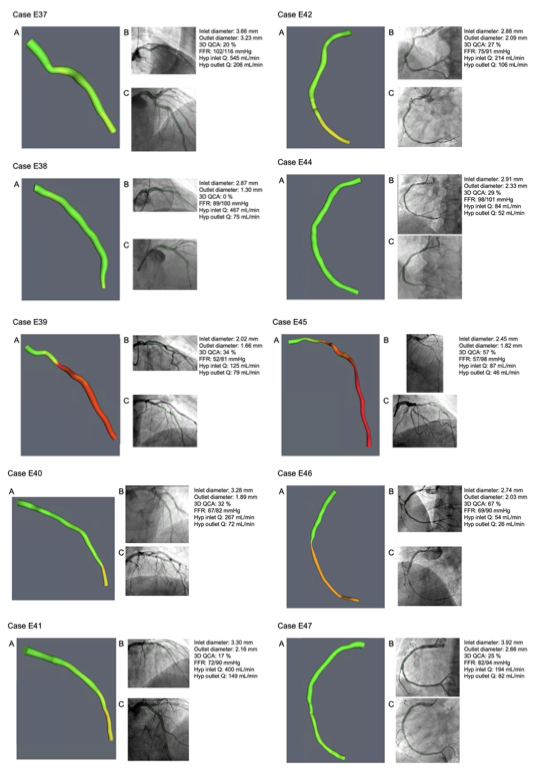

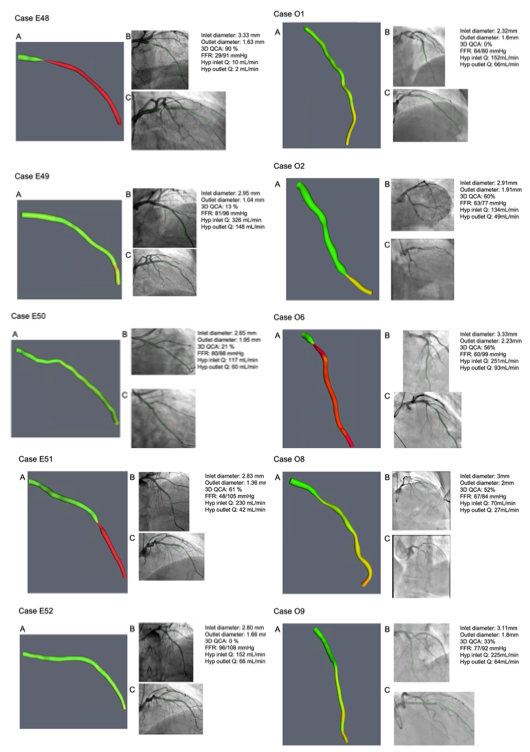

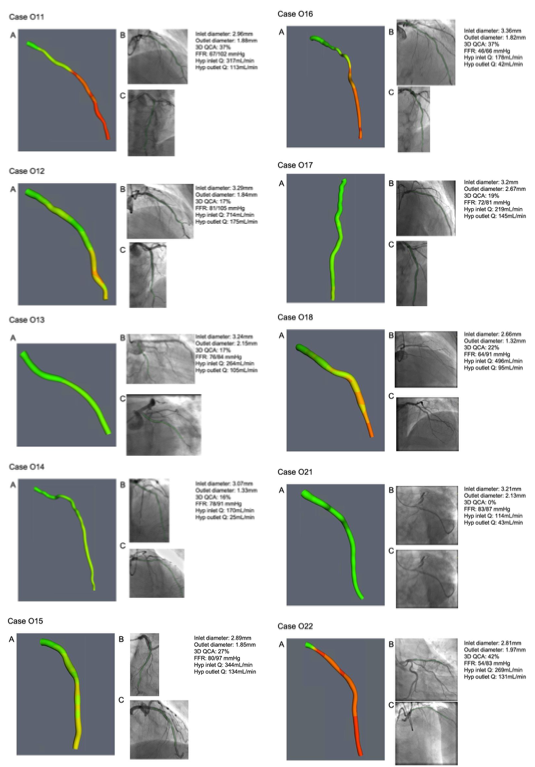

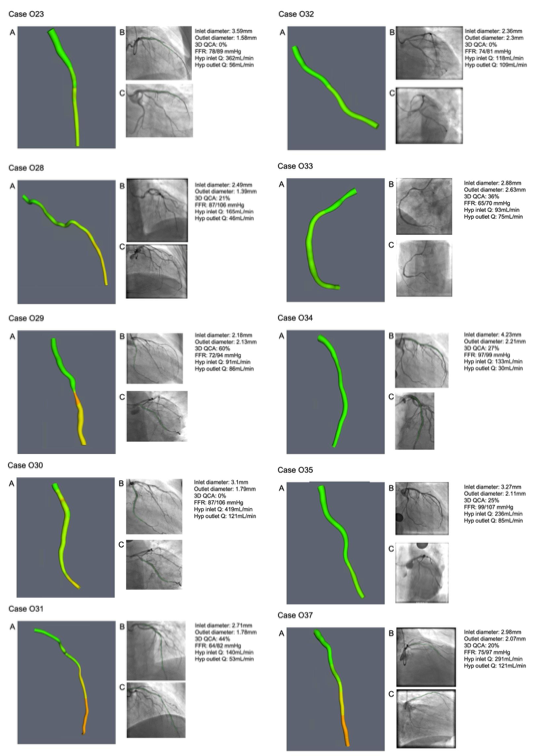

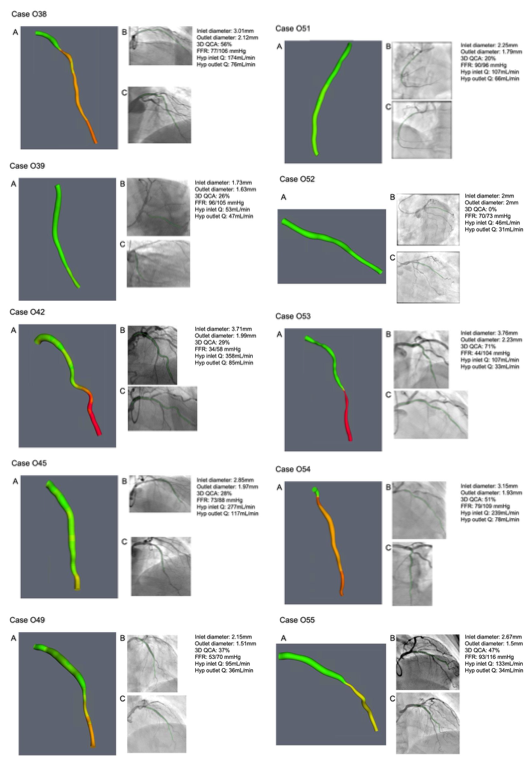

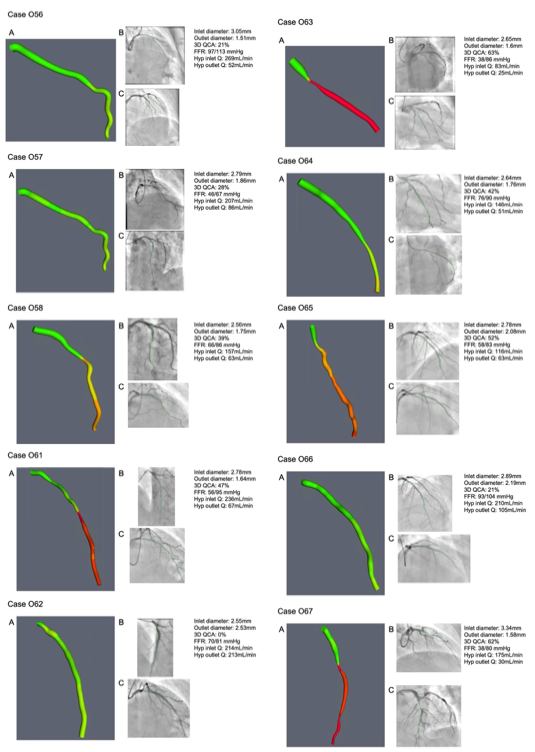

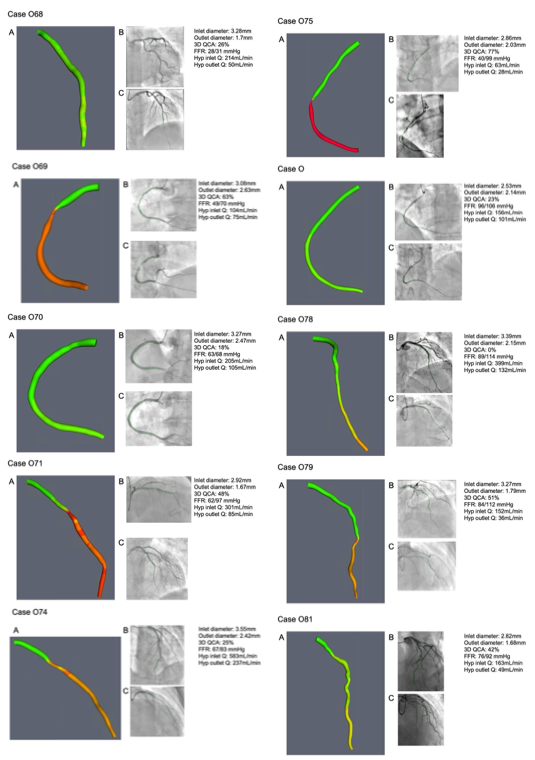

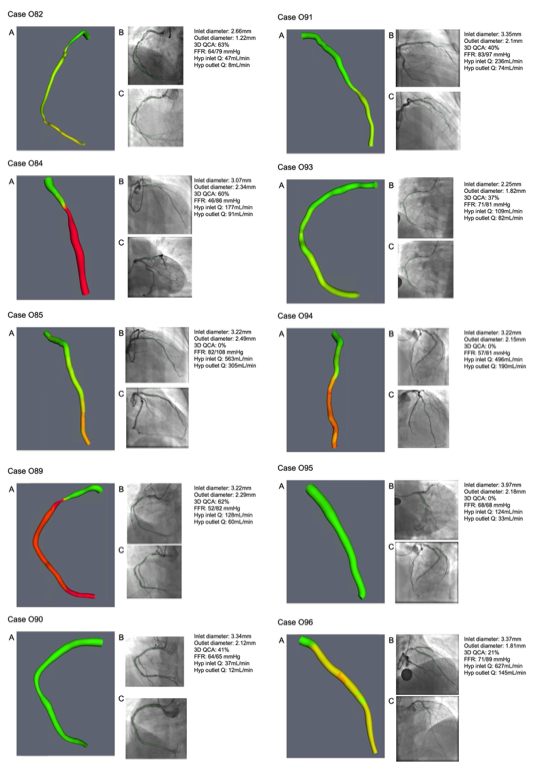

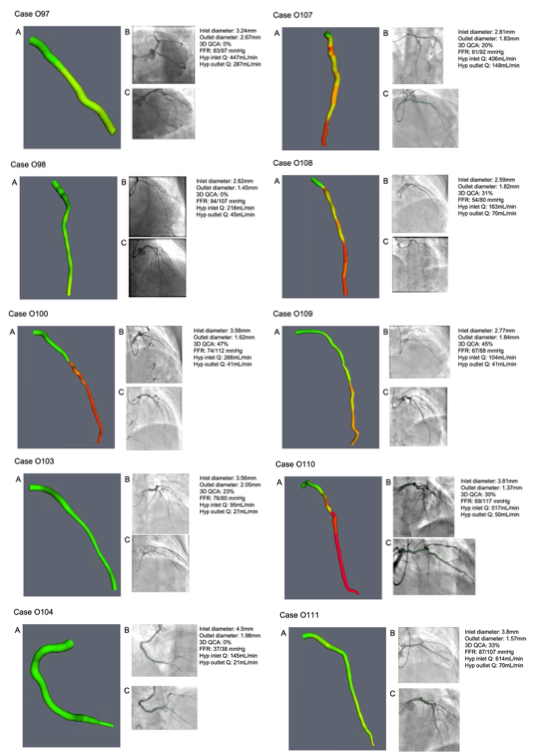

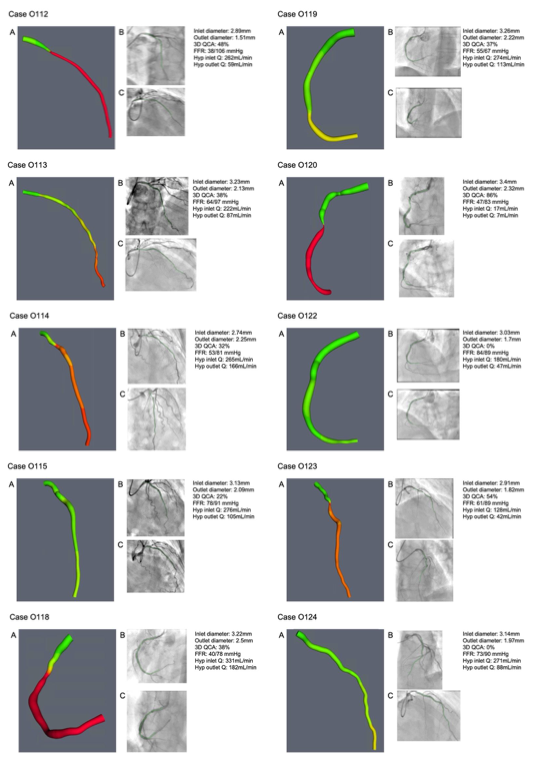

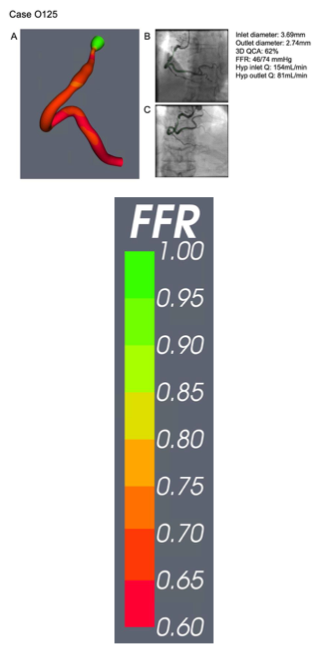

**Demographics and lesion characteristics of excluded patients**

BMI, body mass index; CCS, chronic coronary syndrome; NSTEMI, non-ST elevation myocardial infarction; PCI, percutaneous coronary intervention; STEMI, ST-elevation myocardial infarction.

| **Demographics (n = 13)** | |
| --- | --- |
| Age, y | 67.3 ±12.4 |
| Male | 10 (83%) |
| BMI | 26.4 ±4.3 |
| Current or ex-smoker | 6 (50%) |
| **Enrolment pathology** | |
| CCS | 5 (42%) |
| STEMI | 2 (17%) |
| NSTEMI | 5 (42%) |
| **Comorbidities** | |
| Left ventricular ejection fraction | 51.1 ±8.5 |
| Hypertension | 4 (33%) |
| Diabetes mellitus | 2 (17%) |
| Hypercholesterolaemia | 4 (33%) |
| Previous myocardial infarction | 5 (42%) |
| Previous stroke | 0 |
| **Vessel characteristics (n = 44)** | |
| Left anterior descending | 18 (41%) |
| Left circumflex | 14 (32%) |
| Right coronary artery | 11 (25%) |
| Left internal mammary artery | 1 (2%) |
| Pre-PCI physiology assessment | 23 (59%) |

**Split violin plot for Rayflow (Blue) and virtuQ (Red) assessed MRR**

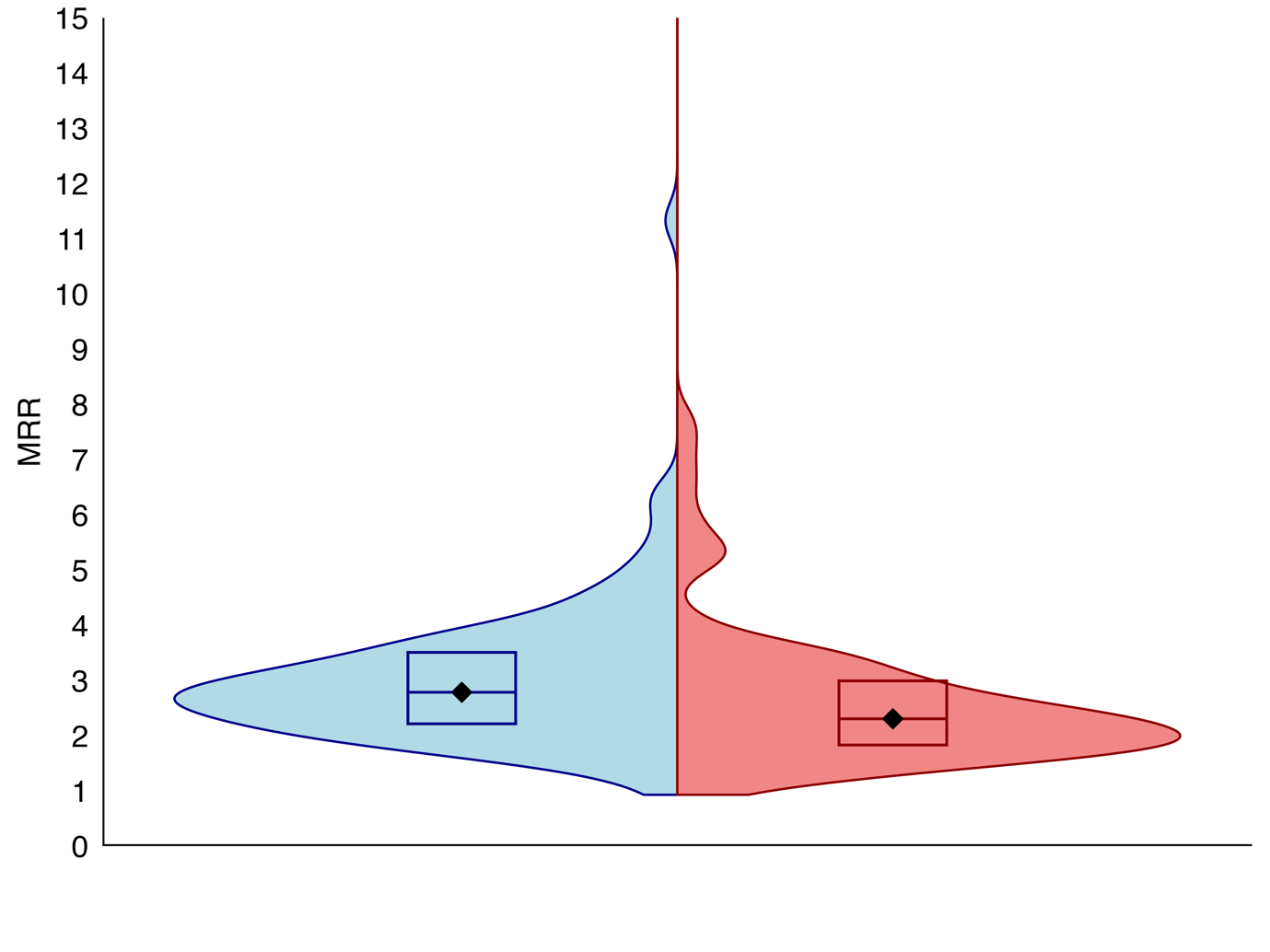

**ROC sensitivity analysis for microvascular resistance reserve (MRR)**

Optimal diagnostic threshold 2.5.

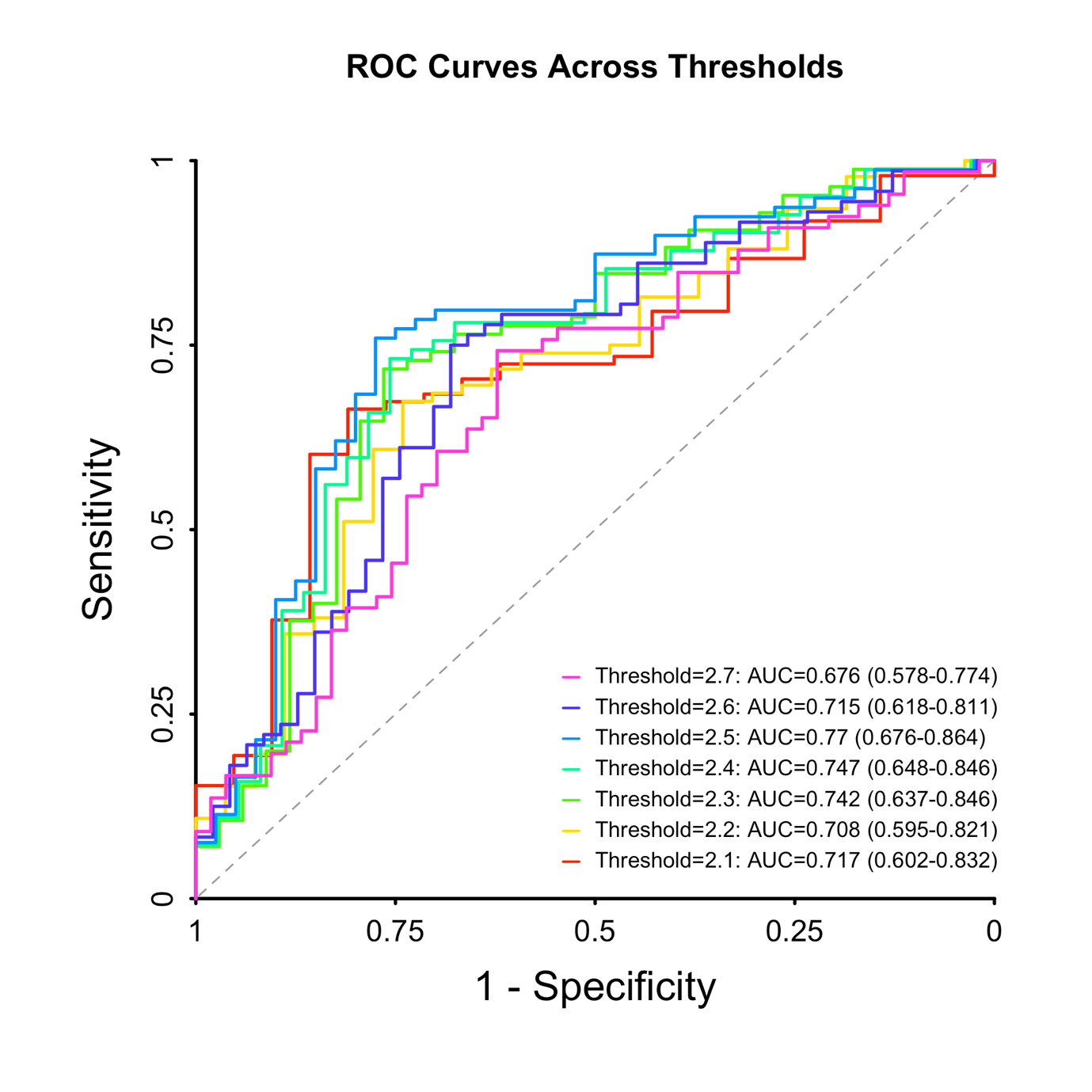

**Alluvial plot for microvascular resistance reserve (MRR)**

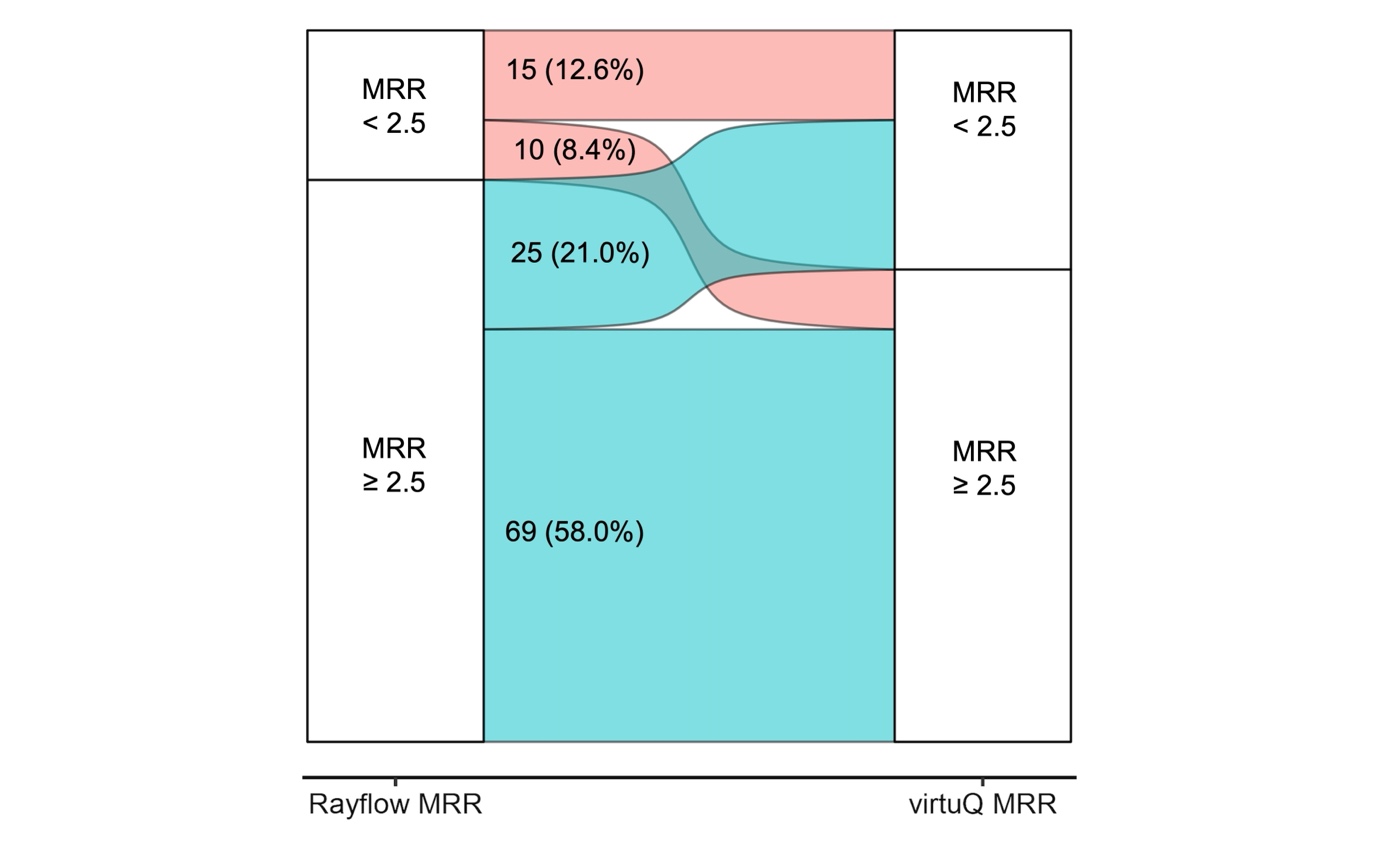

**Quantile regression Bland-Altman plot for microvascular resistance reserve (MRR) displaying regression line equations.**

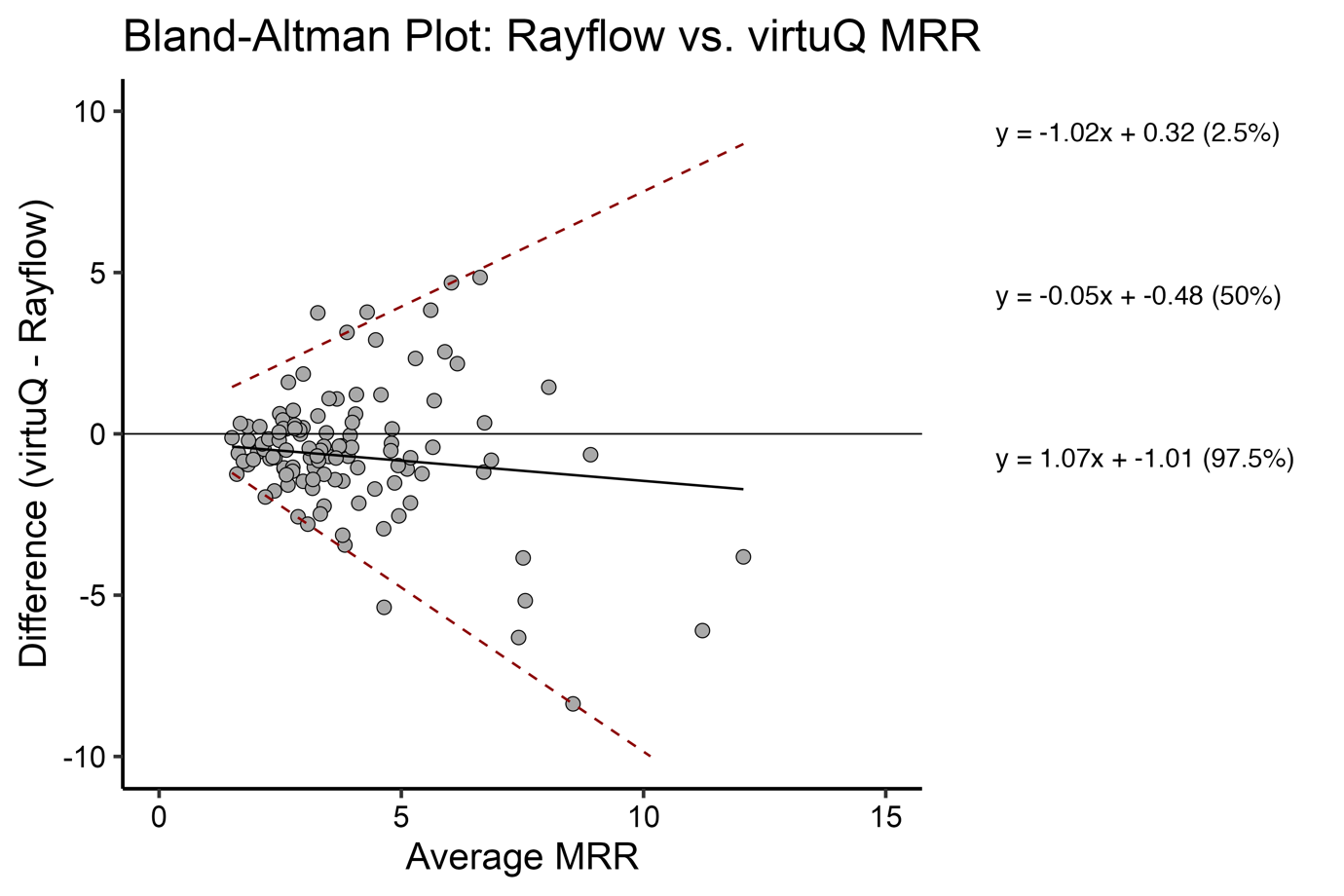

**MRR SHAP analysis results**

Average R^2^ score values for training set (cross-validation train + test) and test set were:

| **Metrics** | **Statistics** | **Random Forest** | **Linear Regression** |
| --- | --- | --- | --- |
| **Cross-validation train R2** | Mean | 0.4607 | 0.3980 |
|  | Median | 0.4589 | 0.3811 |
|  | Standard deviation | 0.0300 | 0.0573 |
| **Cross-validation test R2** | Mean | -0.0157 | 0.0686 |
|  | Median | -0.0051 | 0.0912 |
|  | Standard deviation | 0.1225 | 0.1664 |
| **Test R2 score** | Mean | 0.1553 | 0.1849 |
|  | Median | 0.2020 | 0.2451 |
|  | Standard deviation | 0.2127 | 0.1998 |

These results suggested our MRR data was not suitable for SHAP analysis and was therefore not performed.

**Before and after dot plot for MRR**

Flow-diameter scaling exponent 7/3 (2.33) versus 3.0

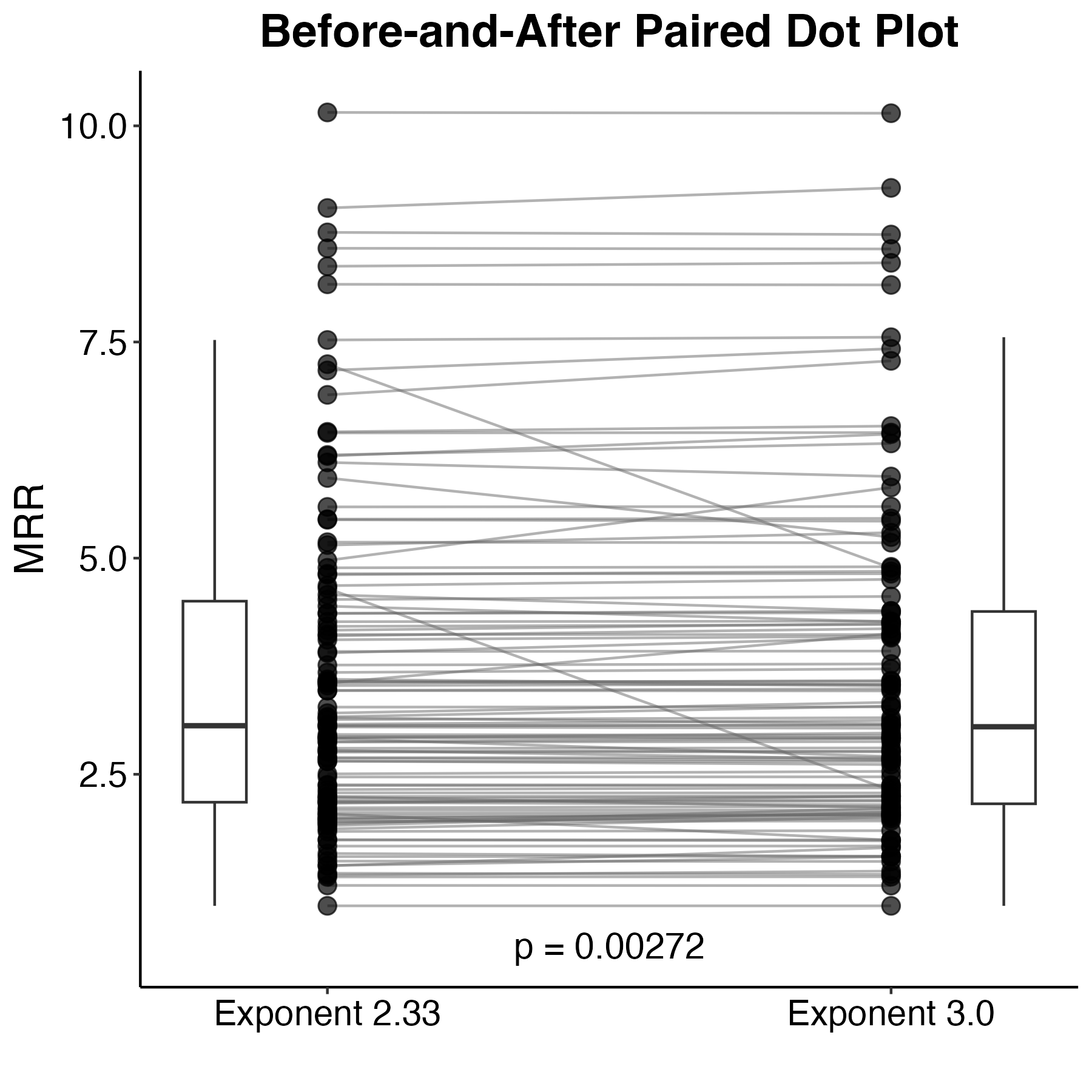

**Split violin plot for Rayflow (Blue) and virtuQ (Red) assessed hyperaemic inlet flow**

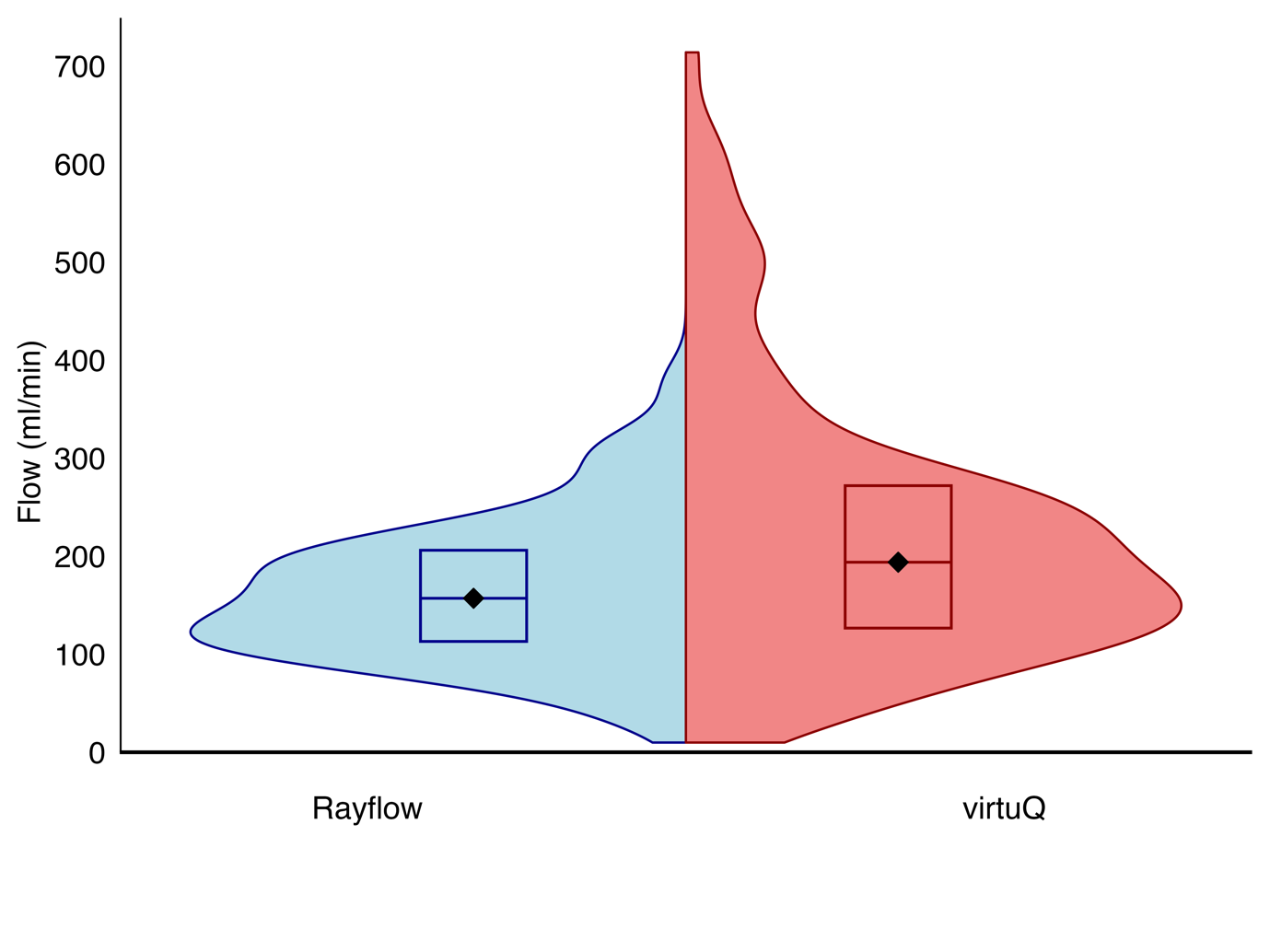

**Rayflow/virtuQ hyperaemic flow Bland-Altman quantile regression line equations**

As seen in main manuscript figure 1.

Q2.5: y = 0.034 * x + -118.37

Q50: y = 0.814 * x + -86.144

Q97.5: y = 1.391 * x + -7.047

**Absolute inlet flow SHAP analysis results**

Average R^2^ score values for training set (cross-validation train + test) and test set were:

| **Metrics** | **Statistics** | **Random Forest** | **Linear Regression** |
| --- | --- | --- | --- |
| **Cross-validation train R^2^** | Mean | 0.6568 | 0.6698 |
|  | Median | 0.6548 | 0.6740 |
|  | Standard deviation | 0.0326 | 0.0217 |
| **Cross-validation test R^2^** | Mean | 0.5277 | 0.5228 |
|  | Median | 0.5351 | 0.5351 |
|  | Standard deviation | 0.0545 | 0.0631 |
| **Test R^2^ score** | Mean | 0.6642 | 0.5668 |
|  | Median | 0.7011 | 0.5995 |
|  | Standard deviation | 0.1580 | 0.1234 |

Averaged SHAP results for Random Forest and Linear Regression in both the test set and fold test sets are shown on the subsequent two pages respectively.

Random forest SHAP analysis for absolute flow:

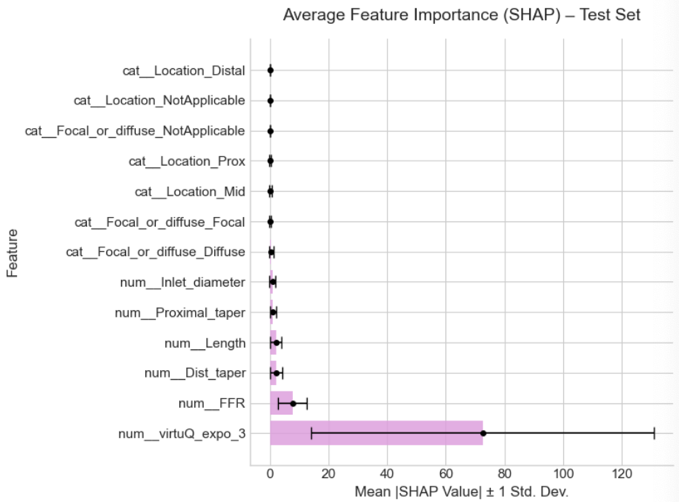

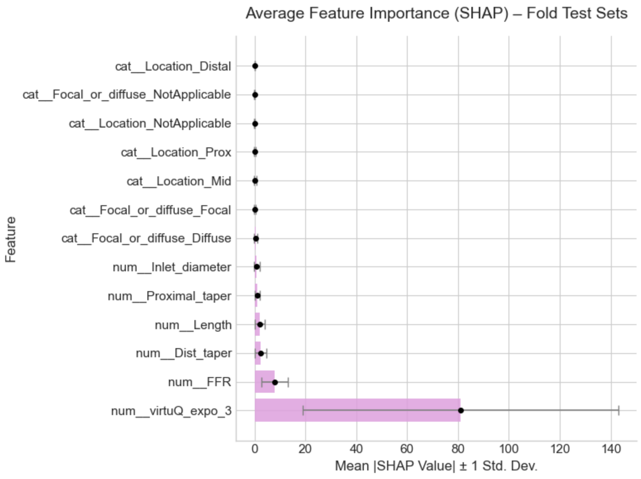

| Number of runs included | 44/60 | | Number of runs included | 44/60 | |
| --- | --- | --- | --- | --- | --- |
| Average score | 0.5537 | | Average score | 0.5537 | |
| SHAP values (Filtered Runs) | | | SHAP values (Filtered Runs) | | |
| Feature | Mean | Std | Feature | Mean | Std |
| num__virtuQ_expo_3 | 72.555 | 58.378 | num__virtuQ_expo_3 | 80.239 | 61.204 |
| num__FFR | 7.702 | 4.954 | num__FFR | 7.689 | 4.769 |
| num__Dist_taper | 2.249 | 2.129 | num__Length | 2.345 | 2.452 |
| num__Length | 2.076 | 2.020 | num__Dist_taper | 1.962 | 2.056 |
| num__Proximal_taper | 1.060 | 0.991 | num__Inlet_diameter | 1.065 | 1.082 |
| num__Inlet_diameter | 0.860 | 1.033 | num__Proximal_taper | 0.750 | 0.676 |
| cat__Focal_or_diffuse_Diffuse | 0.498 | 0.744 | cat__Focal_or_diffuse_Diffuse | 0.369 | 0.513 |
| cat__Focal_or_diffuse_Focal | 0.204 | 0.312 | cat__Location_Mid | 0.243 | 0.317 |
| cat__Location_Mid | 0.194 | 0.408 | cat__Focal_or_diffuse_Focal | 0.178 | 0.234 |
| cat__Location_Prox | 0.152 | 0.236 | cat__Location_Prox | 0.053 | 0.106 |
| cat__Focal_or_diffuse_NA | 0.005 | 0.031 | cat__Location_NA | 0.037 | 0.141 |
| cat__Location_NA | 0.003 | 0.021 | cat__Focal_or_diffuse_NA | 0.000 | 0.000 |
| cat__Location_Distal | 0.000 | 0.000 | cat__Location_Distal | 0.000 | 0.000 |

Ridge Linear Regression SHAP analysis for absolute flow:

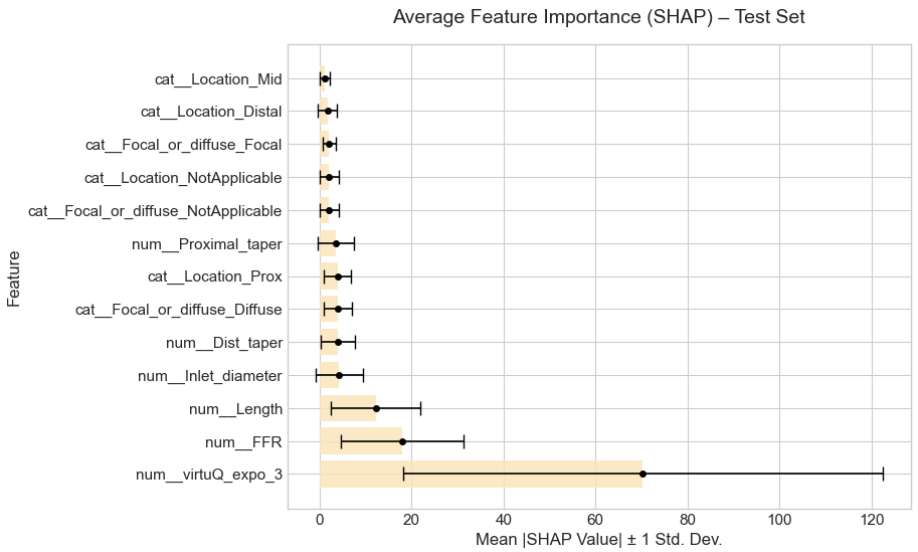

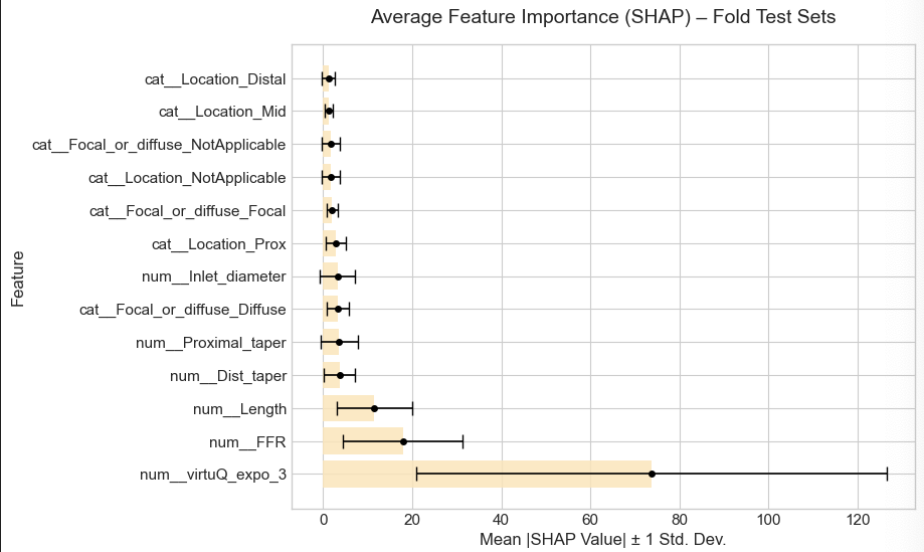

| Number of runs included | 39/60 | | Number of runs included | 39/60 | |
| --- | --- | --- | --- | --- | --- |
| Average score | 0.5617 | | Average score | 0.5617 | |
| SHAP values (Filtered Runs) | | | SHAP values (Filtered Runs) | | |
| Feature | Mean | Std | Feature | Mean | Std |
| num__virtuQ_expo_3 | 70.275 | 52.114 | num__virtuQ_expo_3 | 73.660 | 52.767 |
| num__FFR | 17.921 | 13.415 | num__FFR | 17.964 | 13.421 |
| num__Length | 12.187 | 9.775 | num__Length | 11.537 | 8.506 |
| num__Inlet_diameter | 4.214 | 5.091 | num__Dist_taper | 3.738 | 3.462 |
| num__Dist_taper | 3.958 | 3.736 | num__Proximal_taper | 3.632 | 4.172 |
| cat__Focal_or_diffuse_Diffuse | 3.923 | 2.982 | cat__Focal_or_diffuse_Diffuse | 3.332 | 2.573 |
| cat__Location_Prox | 3.870 | 2.985 | num__Inlet_diameter | 3.266 | 4.003 |
| num__Proximal_taper | 3.462 | 3.902 | cat__Location_Prox | 2.957 | 2.229 |
| cat__Focal_or_diffuse_NA | 2.076 | 2.009 | cat__Focal_or_diffuse_Focal | 2.045 | 1.195 |
| cat__Location_NA | 2.076 | 2.009 | cat__Location_NA | 1.768 | 1.967 |
| cat__Focal_or_diffuse_Focal | 2.076 | 1.424 | cat__Focal_or_diffuse_NA | 1.768 | 1.967 |
| cat__Location_Distal | 1.686 | 2.078 | cat__Location_Mid | 1.252 | 0.897 |
| cat__Location_Mid | 1.191 | 1.085 | cat__Location_Distal | 1.216 | 1.536 |

Despite both methods producing similar results, the Random Forest had greater stability. We therefore concluded these were the more reliable results and thus presented in the main manuscript.

**Before and after dot plot for hyperaemic inlet flow**

Flow-diameter scaling exponent 7/3 (2.33) versus 3.0

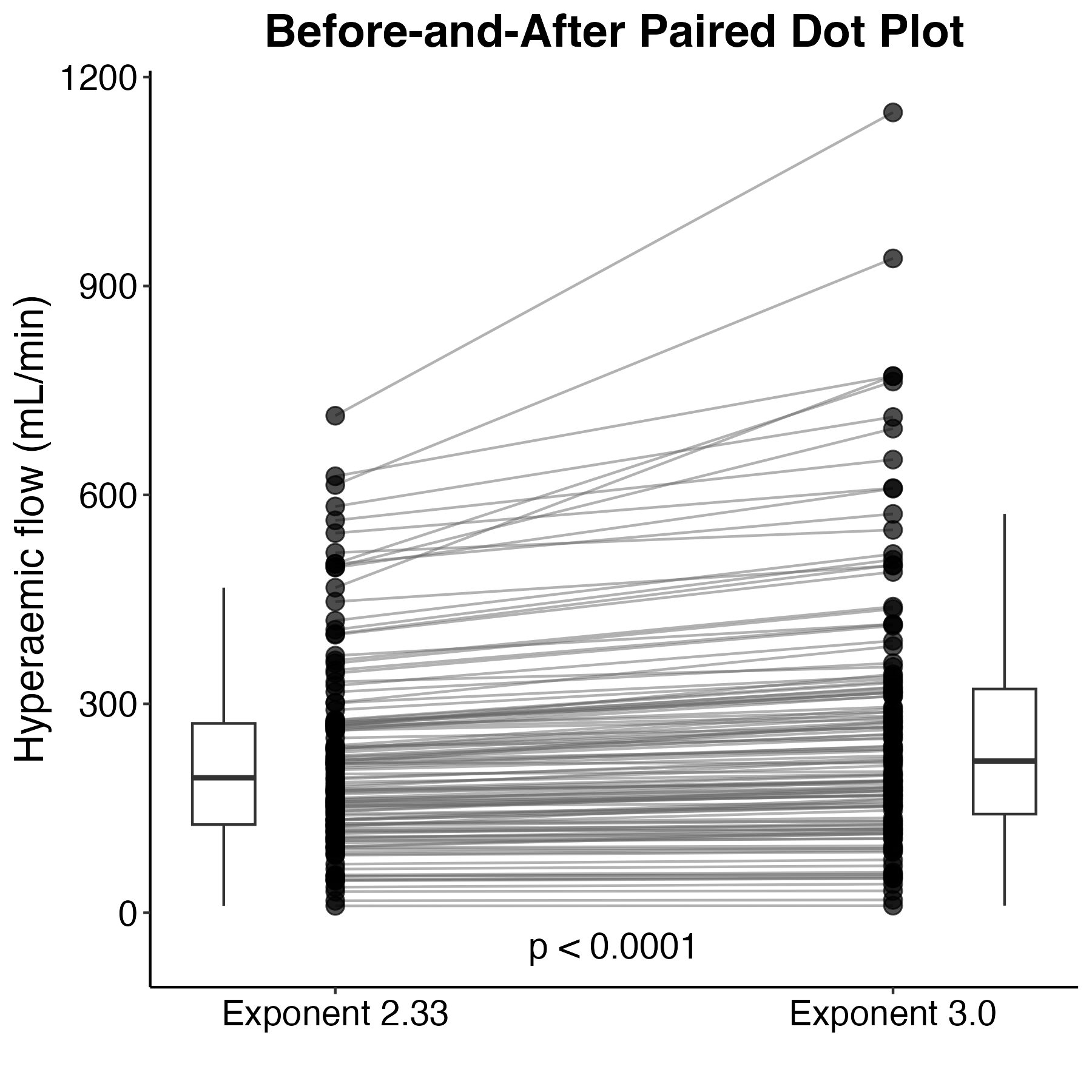

**Summary results for total vessel resistance**

A) Histogram of invasively measured (Rayflow) resistance. B) Histogram of computed (virtuQ) resistance. C) Correlation plot and simple least regression for Rayflow versus virtuQ derived resistance D) Bland Altman plot demonstrating agreement between Rayflow and virtuQ derived resistance. Median bias (solid line, y = 0.69x -321.9) and 95% limits of agreement (dashed lines, y=-1.26x + 11.1 and y=1.75x + 327) derived from quantile regression.

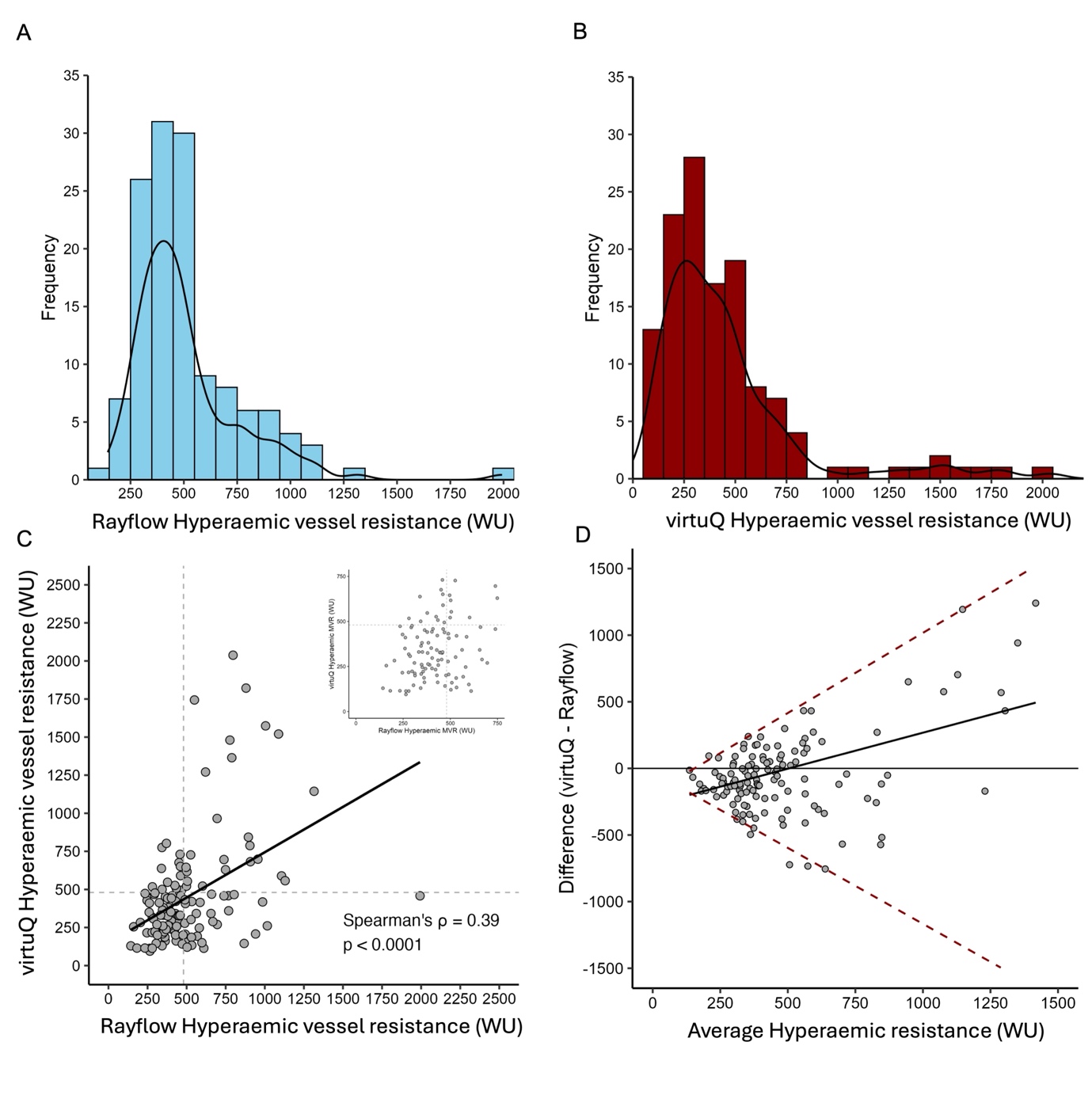

**Summary results for coronary flow reserve (CFR)**

A) Histogram of invasively measured (Rayflow) CFR. B) Histogram of computed (virtuQ) CFR. C) Correlation plot and simple least regression for Rayflow versus virtuQ derived CFR D) Bland Altman plot demonstrating agreement between Rayflow and virtuQ derived CFR. Median bias (solid line, y = 0.44x -1.06) and 95% limits of agreement (dashed lines, y=-1.47x + 1.14 and y=1.21x -1.15) derived from quantile regression. E) Receiver operator characteristic curve for resistance using a diagnostic threshold of 2.5. AUC, area under the curve

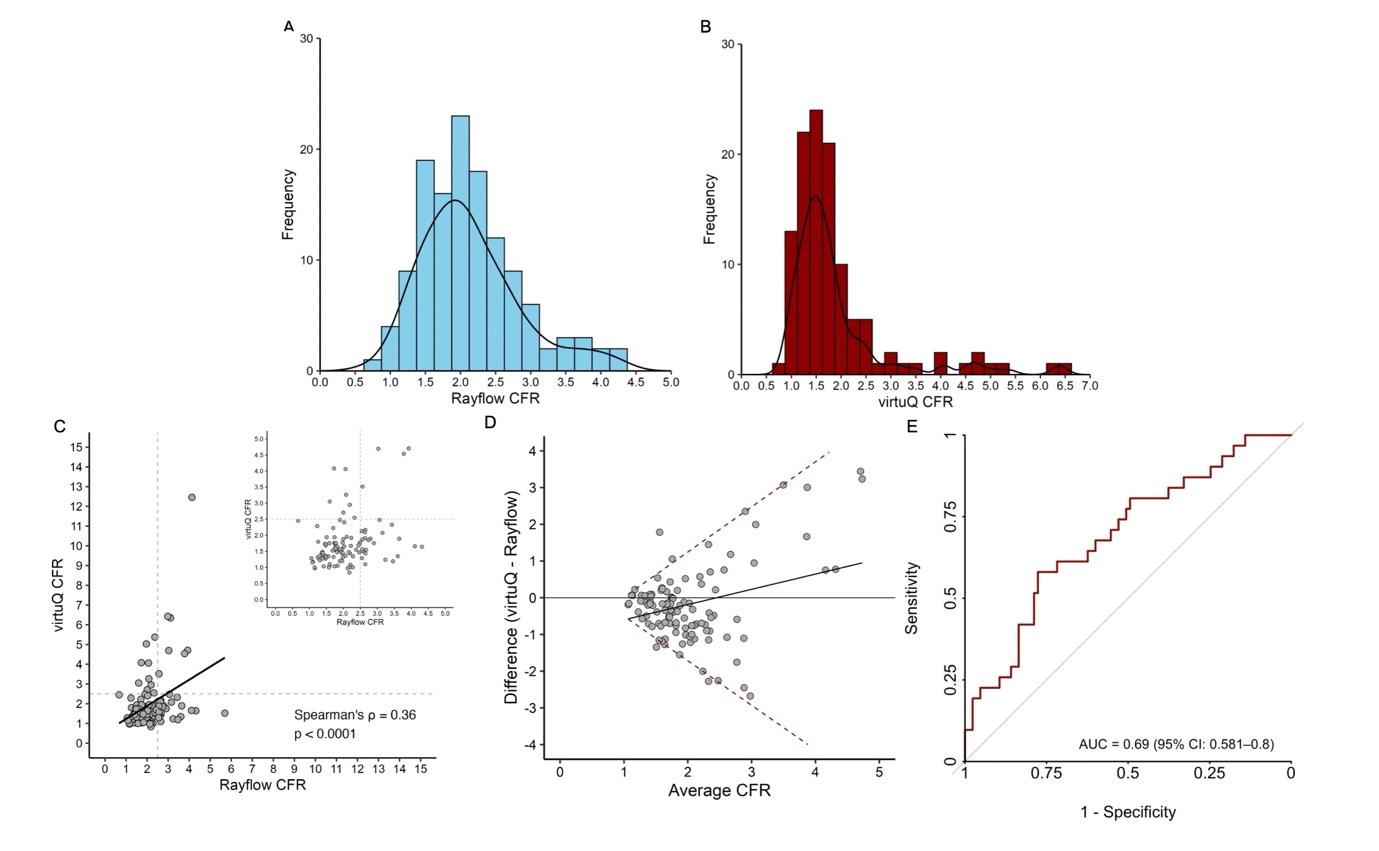

**ROC sensitivity analysis for coronary flow reserve (CFR)**

Optimal diagnostic threshold 2.7.

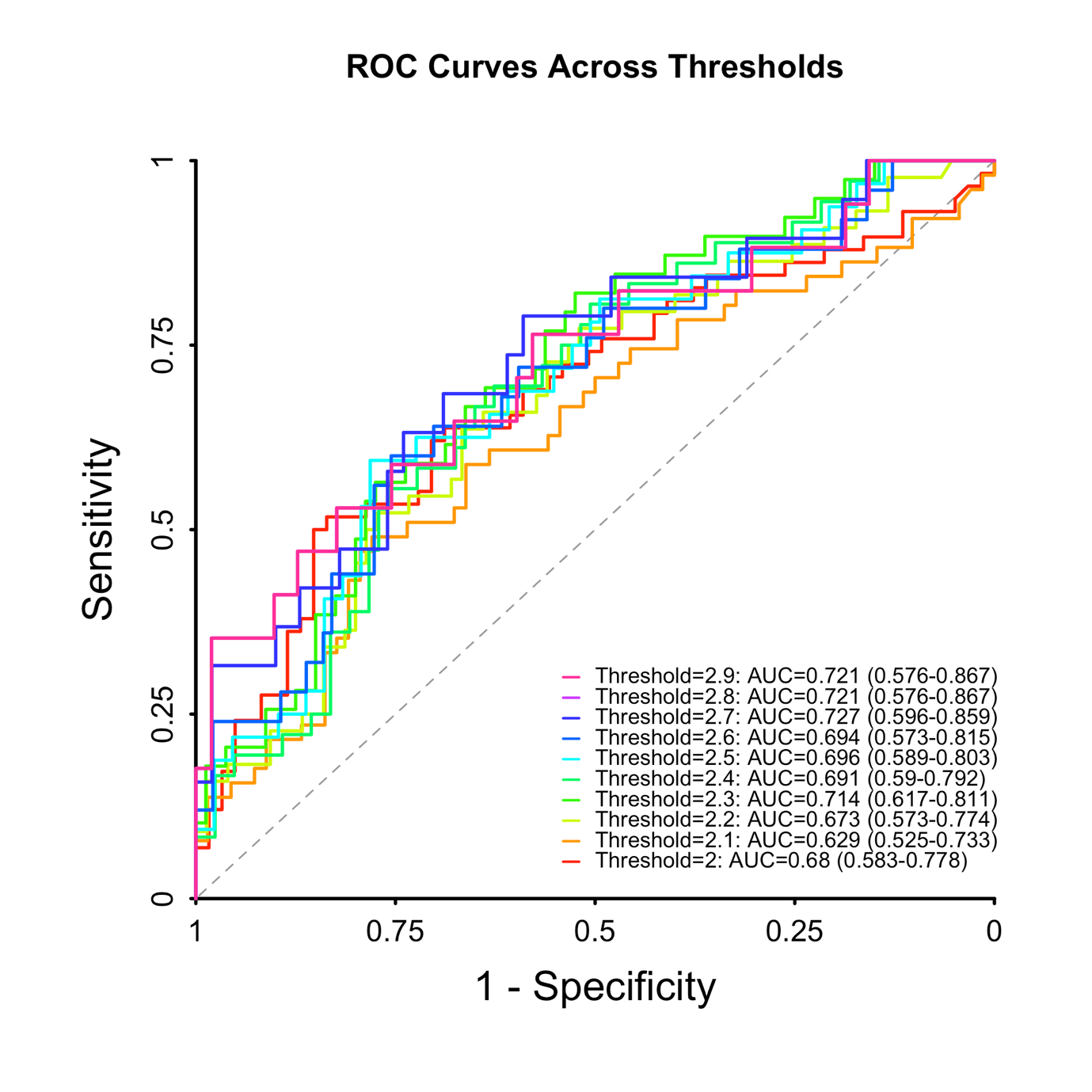
